# To be climate-friendly, food-based dietary guidelines must include limits on total meat consumption – modeling from the case of France

**DOI:** 10.1101/2024.06.10.24308682

**Authors:** Emmanuelle Kesse-Guyot, Julia Baudry, Justine Berlivet, Elie Perraud, Chantal Julia, Mathilde Touvier, Benjamin Allès, Denis Lairon, Serge Hercberg, Hélène Fouillet, Philippe Pointereau, François Mariotti

**Author notes:** Correspondence: Equipe de Recherche en Epidémiologie Nutritionnelle (EREN) SMBH Université Sorbonne Paris Nord, 74 rue Marcel Cachin, 93017 Bobigny, France. **Competing interests:** The authors have no competing interest. Abbreviations: FBDG: Food-based dietary guidelines GBD: Global Burden of Diseases GHGe: greenhouse gas emissions P: percentile TMREL: theoretical minimum-risk exposure level.

## Abstract

Although food-based dietary guidelines (FBDG) include guidelines for meat consumption, their setting most often do not explicitly include environmental considerations. For instance, in France, FBDG recommend consuming no more than 500 g of red meat and 150 g of processed meat per week. This study uses modeling to investigate the range of greenhouse gas emissions (GHGe) that can be achieved under FBDG compliance.

The study analyzed data collected in 2014 from 29,413 NutriNet-Santé participants to assess their adherence to the French FBDG. GHGe, cumulative energy demand (CED), and land occupation (LO) for organic and conventional foods were obtained from the DIALECTE database. Diets adequate in nutrients, culturally acceptable, and consistent with FBDG were modeled under different GHGe constraints. Environmental, nutritional, and health criteria were then calculated.

The average observed adequacy to FBDG was low (19%, SD=25%) and GHGe were 4.34 (SD=2.7%) kgCO2eq/d. The GHGe range of the diets varied from 1.16 to 6.99 kgCO2eq/d, depending up to ∼85% on the level of meat consumption. Similar associations were observed for CED, LO and Health Risk Score. At isoenergetic diets, the diet with the lowest emissions had a higher proportion of vegetables, whole grains, and plant-based substitutes. This diet had a lower CED, LO, and a greater proportion of organic foods when compared to the diet with the highest emissions.

While French dietary guidelines contribute, on average, to mitigating climate change and promoting health, this study emphasizes levers in recommended food consumption to more efficiently reduce diets’ GHGe and points to total meat as the critical issue to better account for pressure on climate change. Other environmental pressures should also be taken into account when designing dietary guidelines.

**Highlights:**

- The average greenhouse gas emissions of the observed diets was 4.34 (SD=2.70) kgCO2eq/d with an energy intake of 2080 Kcal/d
- The diet that closely resembled the observed diet under the dietary guidelines, nutrient and acceptability constraints (TD model) had emissions of 5.15 kgCO2eq/d .
- Modeled diets that complied with dietary guidelines and nutrient and acceptability constraints had emissions ranging from 1.16 kgCO2eq/d (model MinGHGe) to 6.99 kgCO2eq/d (model MaxGHGe).
- All modeled diets had higher consumption levels of fruit, vegetable oils, pulses, and wholegrain products.
- The MinGHGe and MaxGHGe diets, and the range of identified models in between, differed in their level of beef/lamb, refined cereals, fruit, pork, and snack products.
- The level of meat, especially beef/lamb, explained most of the difference (up to ≈85%) in GHGe across models.
- The level of total meat consumption varied progressively across models that imposed an increase in GHGe.

## Introduction

Our food systems are crippling the environment, pushing us beyond critical planetary boundaries and accelerating environmental decline ^1,2^. Food production is a major driver, and six out of nine planetary boundaries have already been breached ^3–5^. For instance, food systems account for 34% of greenhouse gas emissions (GHGe) ^6^ and 70% of blue water usage ^7^. Additionally, land overuse and reliance on synthetic inputs (fertilizers and pesticides) are driving biodiversity loss at an alarming rate ^8^.

Beyond environmental concerns, Diet significantly contributes to the burden of disease ^9,10^. In that context, many countries have developed dietary guidelines in recent decades, with the aim to guide populations towards healthier diets ^11^. However, these guidelines frequently partially account for the profound influence of agriculture and dietary patterns on the environment, despite the intricate interplay between these factors ^11^. Few dietary guidelines were designed while accounting for the goal of minimizing environmental impact when setting consumption targets ^12,13^.

Although there is a wealth of research on the dietary environmental burden associated with adherence to dietary guidelines, environmental impact estimates vary depending on methodological factors. It remains unclear to what extent following dietary guidelines is aligned with reducing environmental pressures. Recently, Springmann *et al.* reported that compliance with most of these official dietary guidelines would yield only a modest 13% reduction in greenhouse gas emissions on average (geographical range: −34% to +35%), as compared to the current situation ^14^.

In France, the French High Council of Public Health (HCSP, Haut Conseil de la Santé Publique) updated national dietary guidelines in 2017 ^15^. The consumption limits recommended by the HCSP were based on scientific literature about the relationships between diet and long-term health and a healthy eating patterns as modelled by The French Food Safety Agency (ANSES) ^16^. This modeling aimed to optimize diets by considering various factors, such as meeting nutrient reference intakes and the established relations betweenconsumption of food groups and long-term health, limiting exposure to certain contaminants, and considering acceptable levels of consumption. Although French dietary guidelines did not explicitly considered environmental pressures when they were implemented, we previously showed that diets closely following these guidelines had an overall reduced environmental footprint compared to non-compliant diets (comparing high versus low adherents: -46% GHGe) ^17^.

However, because this result was based on observed data it does not mean that adherence to dietary guidelines necessarily implies low-emission diets.

FAO and WHO have established a list of 16 principles for sustainable healthy diets ^18^ covering health (8 items), environmental (5 items), and sociocultural (3 items) aspects.

Assessment of the alignment of these principles with existing food-based dietary guidelines (FBDG) has recently been investigated ^11,19,20^. A recent report covering dietary guidelines of 83 countries found that no country addressed all 16 guiding principles across its documents and that FBDG of some countries, such as France, did not fully align with the FAO principles ^11^. Only 45% of FBDG documents mentioned environmental preservation and the vast majority lacked consistency with sustainability.

Therefore, this study aimed to investigate the range of GHGe that could result from diets that follow the French FBDG using optimization modeling. For diets complying with both nutrient and acceptability constraints and with all FBDG individual recommendations, we identified the minimum and maximum GHGe levels. Then, we examined which characteristics of the diets explained the gradual variation in GHGe emissions.

## Materiel & method

### Population

This study was conducted on a sample of adults from the web-based prospective nutritional NutriNet- Santé cohort ^21^. The participants are volunteers recruited from the general French population. This study is conducted in accordance with the Declaration of Helsinki, and all procedures were approved by the Institutional Review Board of the French Institute for Health and Medical Research (IRB Inserm 0000388FWA00005831) and the National Commission on Informatics and Liberty (Commission Nationale de l’Informatique et des Libertés, CNIL 908450 and 909216). Electronic informed consent was obtained from all participants. The NutriNet-Santé study is registered in ClinicalTrials.gov (NCT03335644).

Sociodemographic characteristics, including age, education (<high school diploma, high school diploma, and post-secondary graduate), lifestyles, i.e. smoking status (former, current, or never-smoker) and physical activity level assessed using the International Physical Activity Questionnaire ^22^ as well as anthropometrics data ^23^, are collected using pre-validated questionnaires each year ^24,25^. The participants were asked to report their total monthly income from different sources, such as salary, rental income, family allowance, or social benefits. To determine the monthly household income, the household unit was defined according to the National Institute of Statistics and Economic Studies (INSEE) guidelines ^26^. The first adult in the household was allocated one household unit, while other individuals aged 14 years or older were allocated 0.5 units, and children below 14 years were allocated 0.3 units. We reported data closest to the FFQ (Food Frequency Questionnaire, see below).

### Dietary data

The dietary data were collected in 2014 via a self-administered semi-quantitative FFQ, aiming to distinguish organic (under the official label) and conventional food consumption ^27^. This tool is based on a previously validated 264-item food frequency questionnaire ^28^, improved by a five-point scale to evaluate the mode of production of food ^29^. For each food item, participants reported the frequency of food consumed (over the past 12 months) as organic by ticking the following modalities: “never”, “rarely”, “half-of-time”, “often” or “always” in response to the question ‘How often was the product of organic origin?’. Weight was allocated to each frequency modality, i.e., 0, 25, 50, 75, and 100%, respectively. Nutrient intakes were calculated using a published food composition table ^30^.

### French food-based dietary guidelines and PNNS-GS2

In France, the High Council of Public Health published the revised version of the dietary guidelines for adults in 2017 ^15^, including both specific food consumption targets and general guidelines such as: *“to promote dietary sustainability in the dietary guidelines: opt for raw (unprocessed), seasonal food products, rely on short supply chains and low-input production methods, i.e. with a restriction in inputs”*.

To reflect the level of adherence to these dietary guidelines, a validated dietary score (sPNNS-GS2) has been previously developed and validated ^31^, and showed strong association with a wide range of health outcomes ^31–33^.

The sPNNS-GS2 (theoretical range: -∞ to 14.25), consists of 6 adequacy components and 7 moderation components. The components are weighted according to the level of epidemiological evidence for the associations with health and a penalty on energy intake is also given. if it exceeds nutritional needs. It includes components related to fruits and vegetables, pulses, whole grains, nuts, fish, red meat, processed meat, sweet products, sweet drinks, added lipids, alcohol, dairy products, and salt. Scoring and computation have been extensively described elsewhere ^31^ and are presented in **Supplemental Table 1** and **Supplemental Method 1**.

### Environmental pressure data

Environmental indicators assessment related to food production was computed using life cycle analysis (LCA) using the DIALECTE database developed by Solagro ^34^. GHGe (kg of CO_2_ equivalents (CO_2_eq)), cumulative energy demand (MJ), and land occupation (m^2^) for organic and conventional food production were calculated. Only the production stages have been considered due to a lack of data regarding food production methods for other steps. The packaging, transport, treatment, storage and recycling stages were not included in the scope of the LCA. Extensive details and raw data have been provided elsewhere ^35^.

### Food prices

A database containing the price of each food item was created. The database considers where the food was purchased and the farming method used (organic or conventional). It is based on the Kantar Worldpanel® (40) purchase database, which includes information from 20,000 households and on other data collected from short food-supply chains ^27^.

### Diet modeling

The optimized diets were identified using the procedure SAS/OR ® *optmodel* (version 9.4; SAS Institute, Inc.). A non-linear optimization algorithm with multistart was used to select a solution that is not only a local minimum. The solutions of the optimization procedure provided the consumption in 47 food groups and the % of organic for each of these groups (as the GHGe for a given food group varies depending on the production method). The models’ input parameters were the mean and 95^th^ percentile of the weighted (see below) observed consumption, and the nutrient content of the 47 groups (calculated by weighting the nutritional values of the items constituent of the group by the population consumption of each item). Each group’s GHGe (organic or conventional) were calculated the same way.

### Optimization process and objectives

– First, we identified the modeled diet as closest to the observed diet while complying with all the nutritional, acceptability, and FBDG constraints. This model minimized the total departure (TD) from the observed diet under all these constraints (TD model), according to the formula (1):

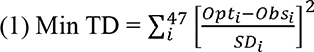

Where opc_i_ and obs_i_ respectively denote the optimized and observed daily consumptions of the food group *i*, with SD*_i_* being its standard deviation in the observed situation.
– Second, we identified the minimum and maximum GHGe values that are compatible with all the nutritional, acceptability, and FBDG constraints, by minimizing or maximizing GHGe under all these constraints, respectively (MinGHGe and MaxGHGe models).
– Finally, we identified a full spectrum of healthy modeled diets of increasing GHGe values (from 1.2 to 6.8 kgCO_2_eq/d) complying with all the nutritional, acceptability, and FBDG constraints, by minimizing the total departure (TD) from the observed diet under all these constraints and an additional constraint on GHGe to cover its whole possible range from its minimum to maximum value, using a grid search by 0.2 kgCO_2_eq/d. GHGe increments.

### Models constraints

#### *Nutrient* constraints

The nutrient constraints, which included daily intakes of energy and a set of nutrients, were based on the lower and/or upper ANSES 2016 dietary reference intakes ^36^. A specific constraint imposed energy intake to be between +8% and -8% of energy requirements. Lower bounds were defined as either recommended dietary allowance (population reference intake), adequate intake, or lower bound of reference range for the intake in the French population ^36^. Upper bounds were defined as the maximum tolerable intakes for vitamins and minerals or the upper limit of the reference intake range. For zinc and iron, bioavailability was considered using validated equations (**Supplemental Method 2 and Supplemental Method 3**) ^37,38^. A minor acceptable lowest limit than nutritional references based on deficiency intake has been defined for bioavailable zinc and iron as previously published ^39^. The lower threshold values used in this context pertain to a deficiency prevalence of <5%. This approach offers greater flexibility in identifying diets that are considered healthier overall despite the higher prevalence of iron deficiency anemia ^39^. All reference values used are shown in **Supplemental Table 2**.

Of note, the French nutritional reference values for adults are based on the specific physiological requirements of males and females, and established separately for each gender ^36^. Additionally, the reference values are further differentiated for females based on their iron requirements, with a distinction made between females with high and low/moderate iron requirements. To create new nutritional reference values more representative of the average individual, we have derived a weighted average of requirements for males, females with high iron requirements, and females with low/moderate iron requirements. Therefore, the reference values for each nutrient for this average individual are defined as the weighted average requirements of males and females (**Supplemental Table 2**). For adequate intake, based on observed mean intake, the lower limit was set at the 5^th^ weighted percentile value of the overall population.

#### FGDB constraints

To comply with official French dietary guidelines ^15^, models were additionally constrained on the consumption of different food groups using thresholds set by the official French FBDG quantitatively translated as for the PNNS-GS computation (see **Supplemental Table 1**).

– Consumption of fruit and vegetables (including 100% fruit juice up to a maximum of one portion) ≥ 400g/d
– Consumption of 100%fruit juice ≤ 150g/d
– Consumption of nuts ≥ 30g/d
– Consumption of pulses ≥ 400g/wk (i.e. ≥57g/d)
– Consumption of dairy products: 2 portions/d (with a portion for milk=150mL, cheese=30g, yogurt=125g, “*Petits Suisses*”=120g, cottage cheese=100g)
– Consumption of wholegrain products ≥400g/wk (i.e. ≥57g/d)
– Consumption of red meat ≤ 500g/wk (i.e. ≤71.4g/d)
– Consumption of processed meat ≤ 150g/wk (i.e. ≤21.4g/d)
– Consumption of total seafood 2 portions/wk (with a portion=100g, i.e. 28.57g/d)
– Consumption of fatty fish 2 portions/wk (with a portion=100g, i.e. 14.28g/d)
– Consumption of added fat (added lipids) ≤16% of total energy intake
– Consumption of sweet drinks (including other fruit juice, sweet and artificially sweetened beverages) = 0

Of note, no constraints were imposed on salt and sugary foods, as the limitations of sodium and sugar were already considered by the nutrient-related constraints.

#### Acceptability constraints

In order to prevent the models from giving aberrant values (i.e. excessive intakes for some food groups), the maximum possible intake for each food group was set at the 99^th^ percentile of observed consumption distribution. For cereals (refined and wholegrain),a so-called “coupling” limit allowed inter-group substitution so that each can exceed its 99^th^ percentile while only the sum was constrained to the 99^th^ percentile. In addition, as pulses consumption is very low in the observed diet, no acceptability constraint was used for this food group.

#### Sensitivity analyses

Two sensititvity analyses were conducted. First, a sensitivity analysis was also conducted to compare the results under different FBDG constraints to identify the changes in the maximum GHGe values when further restricting the total meat consumption, from 500 (main scenario) to 400, 300, or 200g/wk. Then, the full spectrum of healthy modeled diets of increasing imposed GHGe values complying with nutritional, acceptability, and FBDG constraints, by minimizing the total departure (TD) was reanalyzed with the use of the 95^th^ percentile of the food group consumption distribution as acceptability criteria.

### Descriptive statistics

The observed situation was based on NutriNet-Santé participant data who had completed the FFQ between June and December 2014 (N=37,685), with no missing covariates (N=37,305), who were not under or over-energy reporters (N=35,196), living in mainland France (N=34,453), and with information as regards the place of purchase for the computation of the dietary monetary cost (N= 29,413). Observed sociodemographic, and lifestyle characteristics of the sample were estimated as mean (SD) or percentage according to sex-specific quintiles of the PNNS-GS2.

The modeled diets were described in terms of food group consumption (the 47 food groups used for optimization were grouped into 25 groups for clarity purposes, see **Supplemental Table 4**), nutrient intakes, potential health risk, assessed using the Health Risk Score (HRS), compared to the theoretical maximal risk exposure level (TMREL) of the 2019 Global Burden of Diseases (GBD) study, environmental pressures (GHGe, CED, and LO) as well as monetary cost of the diets. The HRS is presented in **Supplementary Method 4.**

All statistical analyses were performed using SAS® (version 9.4; SAS Institute, Inc., Cary, NC, USA) and Figures were developed using R version 3.6.

## Results

### Observed diets

In the observed situation, the weighted mean (SD) age was 55 years (14), and sPNNS-GS2 was 2.28 (3.57). The average GHGe was 4.34 ± 2.70 kgCO2eq/d (at the farm perimeter) (**Table 1**). The sample characteristics by sPNNS-GS2 quintiles are presented in **Supplemental Table 3**. Better adherence to dietary guidelines was associated with older age and higher levels of education, income, and physical activity. A negative association was observed for smoking and living with a partner. Adherence was negatively associated with daily energy intake, but positively associated with consumption of organic foods and the proportion of plant protein in total protein intake.

**Table 1:**
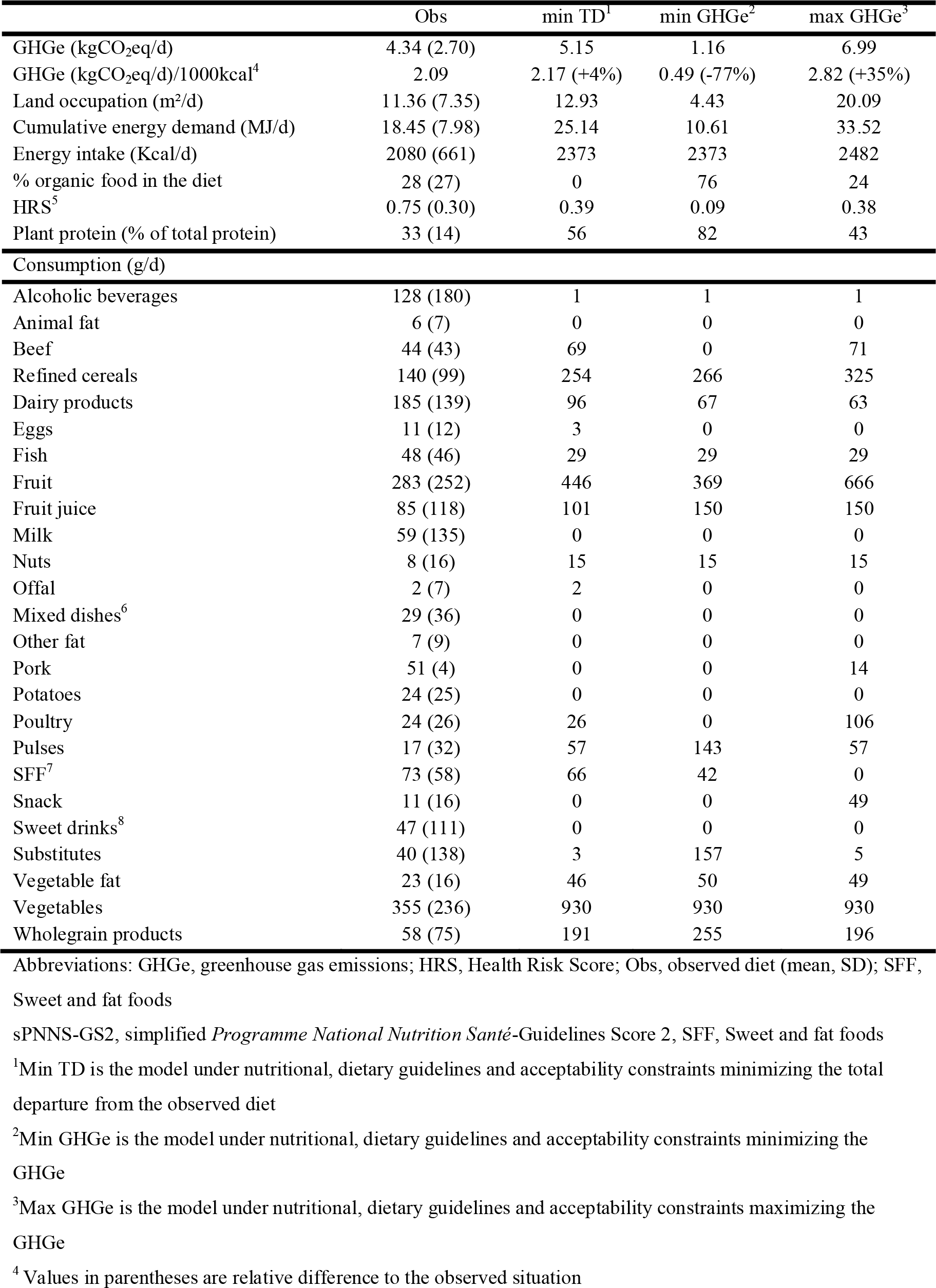
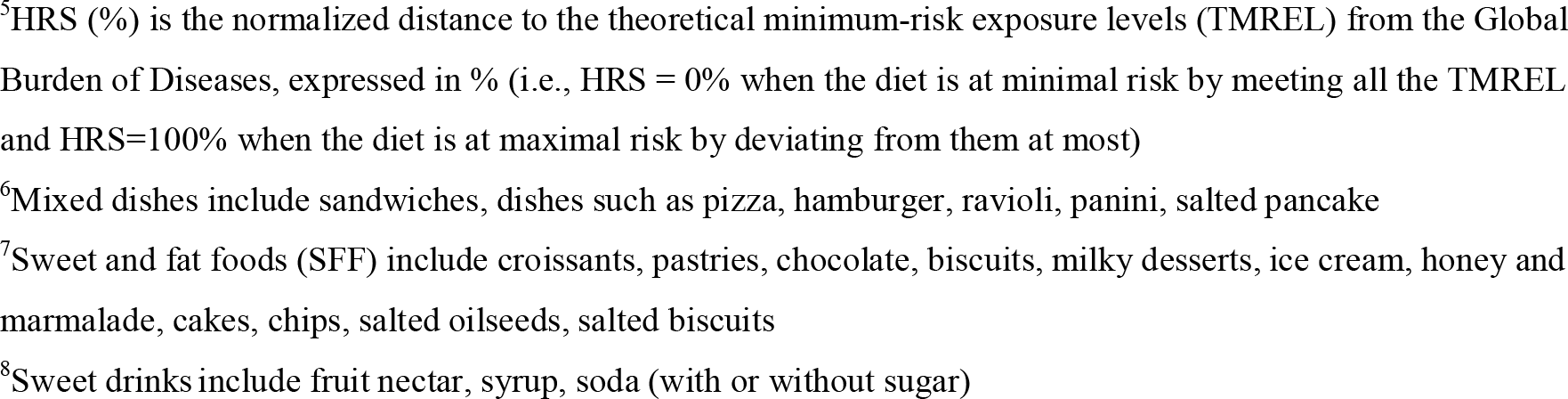
Characteristics of observed diets and main modeled diets.

|  | Obs | min TD<sup>1</sup> | min GHGe<sup>2</sup> | max GHGe<sup>3</sup> |
| --- | --- | --- | --- | --- |
| GHGe (kgCO<sub>2</sub>eq/d) | 4.34 (2.70) | 5.15 | 1.16 | 6.99 |
| GHGe (kgCO<sub>2</sub>eq/d)/1000kcal<sup>4</sup> | 2.09 | 2.17 (+4%) | 0.49 (-77%) | 2.82 (+35%) |
| Land occupation (m<sup>2</sup>/d) | 11.36 (7.35) | 12.93 | 4.43 | 20.09 |
| Cumulative energy demand (MJ/d) | 18.45 (7.98) | 25.14 | 10.61 | 33.52 |
| Energy intake (Kcal/d) | 2080 (661) | 2373 | 2373 | 2482 |
| % organic food in the diet | 28 (27) | 0 | 76 | 24 |
| HRS<sup>5</sup> | 0.75 (0.30) | 0.39 | 0.09 | 0.38 |
| Plant protein (% of total protein) | 33 (14) | 56 | 82 | 43 |
| Consumption (g/d) |  |  |  |  |
| Alcoholic beverages | 128 (180) | 1 | 1 | 1 |
| Animal fat | 6 (7) | 0 | 0 | 0 |
| Beef | 44 (43) | 69 | 0 | 71 |
| Refined cereals | 140 (99) | 254 | 266 | 325 |
| Dairy products | 185 (139) | 96 | 67 | 63 |
| Eggs | 11 (12) | 3 | 0 | 0 |
| Fish | 48 (46) | 29 | 29 | 29 |
| Fruit | 283 (252) | 446 | 369 | 666 |
| Fruit juice | 85 (118) | 101 | 150 | 150 |
| Milk | 59 (135) | 0 | 0 | 0 |
| Nuts | 8 (16) | 15 | 15 | 15 |
| Offal | 2 (7) | 2 | 0 | 0 |
| Mixed dishes<sup>6</sup> | 29 (36) | 0 | 0 | 0 |
| Other fat | 7 (9) | 0 | 0 | 0 |
| Pork | 51 (4) | 0 | 0 | 14 |
| Potatoes | 24 (25) | 0 | 0 | 0 |
| Poultry | 24 (26) | 26 | 0 | 106 |
| Pulses | 17 (32) | 57 | 143 | 57 |
| SFF<sup>7</sup> | 73 (58) | 66 | 42 | 0 |
| Snack | 11 (16) | 0 | 0 | 49 |
| Sweet drinks<sup>8</sup> | 47 (111) | 0 | 0 | 0 |
| Substitutes | 40 (138) | 3 | 157 | 5 |
| Vegetable fat | 23 (16) | 46 | 50 | 49 |
| Vegetables | 355 (236) | 930 | 930 | 930 |
| Wholegrain products | 58 (75) | 191 | 255 | 196 |

Abbreviations: GHGe, greenhouse gas emissions; HRS, Health Risk Score; Obs, observed diet (mean, SD); SFF,
Sweet and fat foods
sPNNS-GS2, simplified *Programme National Nutrition Santé*-Guidelines Score 2, SFF, Sweet and fat foods
<sup>1</sup>Min TD is the model under nutritional, dietary guidelines and acceptability constraints minimizing the total departure from the observed diet
<sup>2</sup>Min GHGe is the model under nutritional, dietary guidelines and acceptability constraints minimizing the GHGe
<sup>3</sup>Max GHGe is the model under nutritional, dietary guidelines and acceptability constraints maximizing the GHGe
<sup>4</sup> Values in parentheses are relative difference to the observed situation
<sup>5</sup>HRS (%) is the normalized distance to the theoretical minimum-risk exposure levels (TMREL) from the Global Burden of Diseases, expressed in % (i.e., HRS = 0% when the diet is at minimal risk by meeting all the TMREL and HRS=100% when the diet is at maximal risk by deviating from them at most) <sup>6</sup>Mixed dishes include sandwiches, dishes such as pizza, hamburger, ravioli, panini, salted pancake <sup>7</sup>Sweet and fat foods (SFF) include croissants, pastries, chocolate, biscuits, milky desserts, ice cream, honey and marmalade, cakes, chips, salted oilseeds, salted biscuits <sup>8</sup>Sweet drinks include fruit nectar, syrup, soda (with or without sugar)

Participants in the Q5 had higher or much higher consumption of plant products, especially fruits and vegetables, oilseeds, pulses, whole grains, and plant substitutes, compared with individuals in Q1.

Higher adherence was associated with higher GHGe, even after adjusting for energy intake.

### Modeled diets

When modeling a diet under nutritional constraints and PNNS guidelines, without constraints on GHGe, emissions increased to 5.15 kgCO_2_eq/d (model TD, i.e. as closely as possible to the observed diet), i.e. +4%/1000 kcal compared to the observed diet (**Table 1**). Diets that complied with nutritional, acceptability constraints, and dietary guidelines, had emissions ranging from 1.16 kgCO_2_eq/d (model MinGHGe) to 6.99 kgCO_2_eq/d (model MaxGHGe) (**Table 1**), i.e. -76.7 to +34.8%/1000 kcal compared to the observed diet.

Similar results were observed for LO and CED. The TD model contained no organic food (as by construction, no constraints depending on the mode of production were introduced to the model), while from Min to MaxGHGE models, %organic food products varied from 24%(Max) to 76%(Min) . In the TD model (Table 1), certain food items such as alcoholic beverages, animal fats, milk, other fats, pork, potatoes, snack foods, and soft drinks were excluded compared to the observed diet due to the nutritional and FBDG constraints. Of note, the total meat intake (beef/lamb, poultry) in the TD model was high, 97g/d, far above the target value of the FBDG (500g/wk).

The same foods were also eliminated in both the MinGHGe and MaxGHGe models. For all three models (i.e. TD, MinGHGe and MaxGHGe), there was a systematic increase, compared to the observed diet, of the consumption of fruits, fruit juices, vegetable oil, pulses, vegetable oils, and wholegrain products. Conversely, consumptions of eggs, fish, dairy products, and fatty and sweet products were reduced.The MinGHGe and MaxGHGe models differed in their level of beef/lamb, refined cereals, fruit, pork, and snack products, for which we saw an increase in consumption in the MaxGHGe model, while pulses, wholegrain products, and plant-based substitutes (especially soya-based products) experienced a decrease. Notably, there was a progressive shift towards plant-based diets from the MaxGHGe to MinGHGe models, as expressed by the higher % of protein derived from plant sources from 43% to 82%.

Figure 1 describes various indicators for the GHGe-imposed diets. Specifically, higher GHGe correlated with increases in other environmental indicators such as LO and CED. Similarly, the price of the modeled diets and their HRS increased with GHGe. Conversely, the proportion of organic food in the diet increased non-linearly and then fell drastically. Additionally, the distance from the observed diet exhibited a U-shaped curve, with the levels furthest from the observed diet found at low and high GHGe extremes.

**Figure 1:**
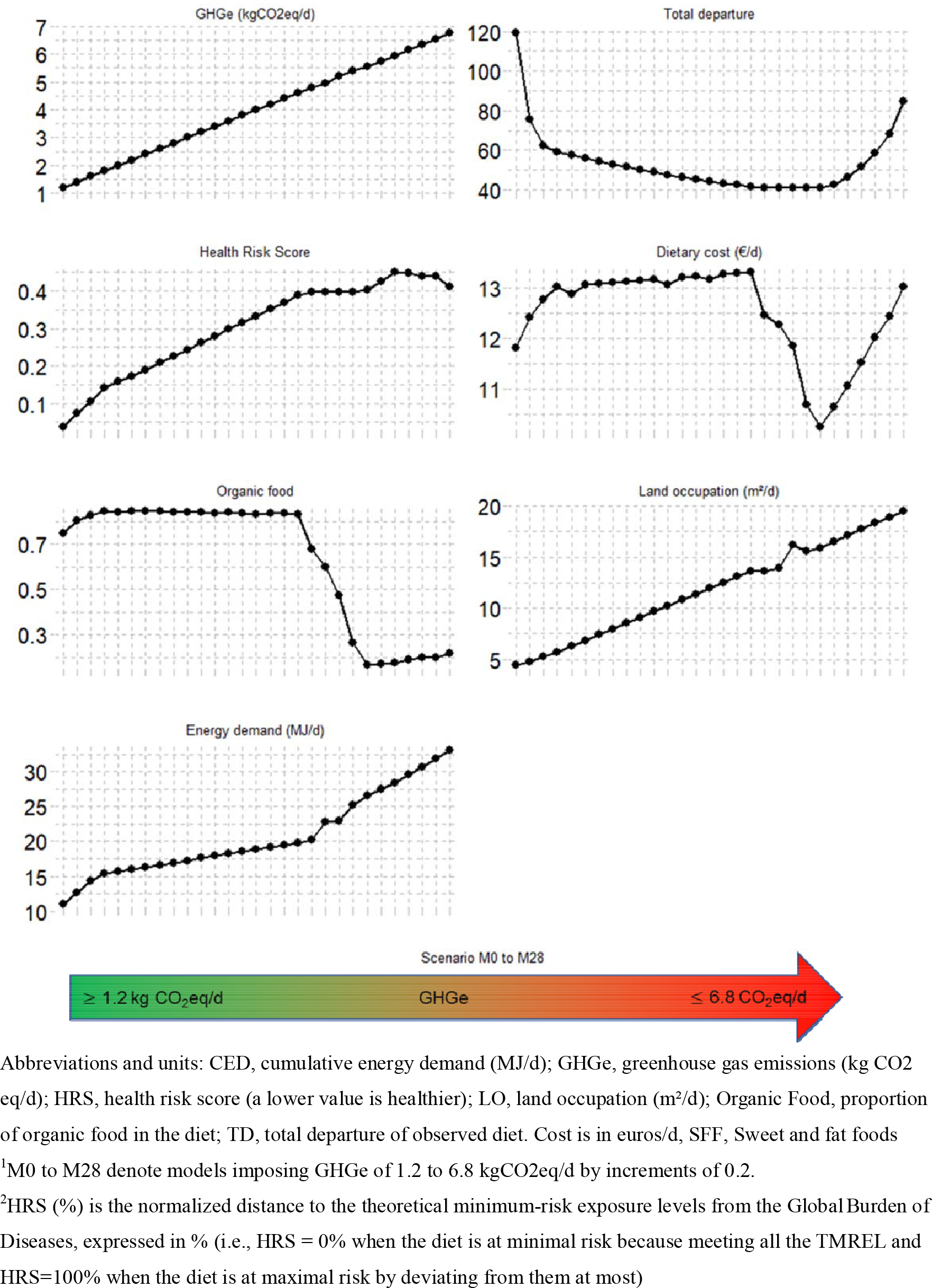
Characteristics of modeled diets adhering to dietary guidelines with minimal deviation from observed diets at different levels of GHGe ^1–2^

The composition of the GHGe-imposed modeled diets is shown in Figure 2 and **Supplemental Table 4**. A gradual increase in GHGe was associated with progressive variations in most types of consumption. Notably, the increase in GHGe was associated with beef/lamb consumption increase, along with a reduction of fruit juices and poultry while pulses and plant-based substitutes. In addition, a slight decrease in the consumption of wholegrain cereals and an increase in refined cereals were observed. Vegetable consumption remained stable, as did fish consumption. Some foods, such as dairy products, offals, and sweet and fatty foods, exhibited a bell-shaped distribution. Finally, vegetables, fish, and oilseeds were found at the upper or lower limits defined in the models. Certain foods like animal fats, eggs, and potatoes were excluded from the modeled diets. Pork consumption did not display a discernible pattern but was most prevalent in the diet with the highest emissions.

**Figure 2:**
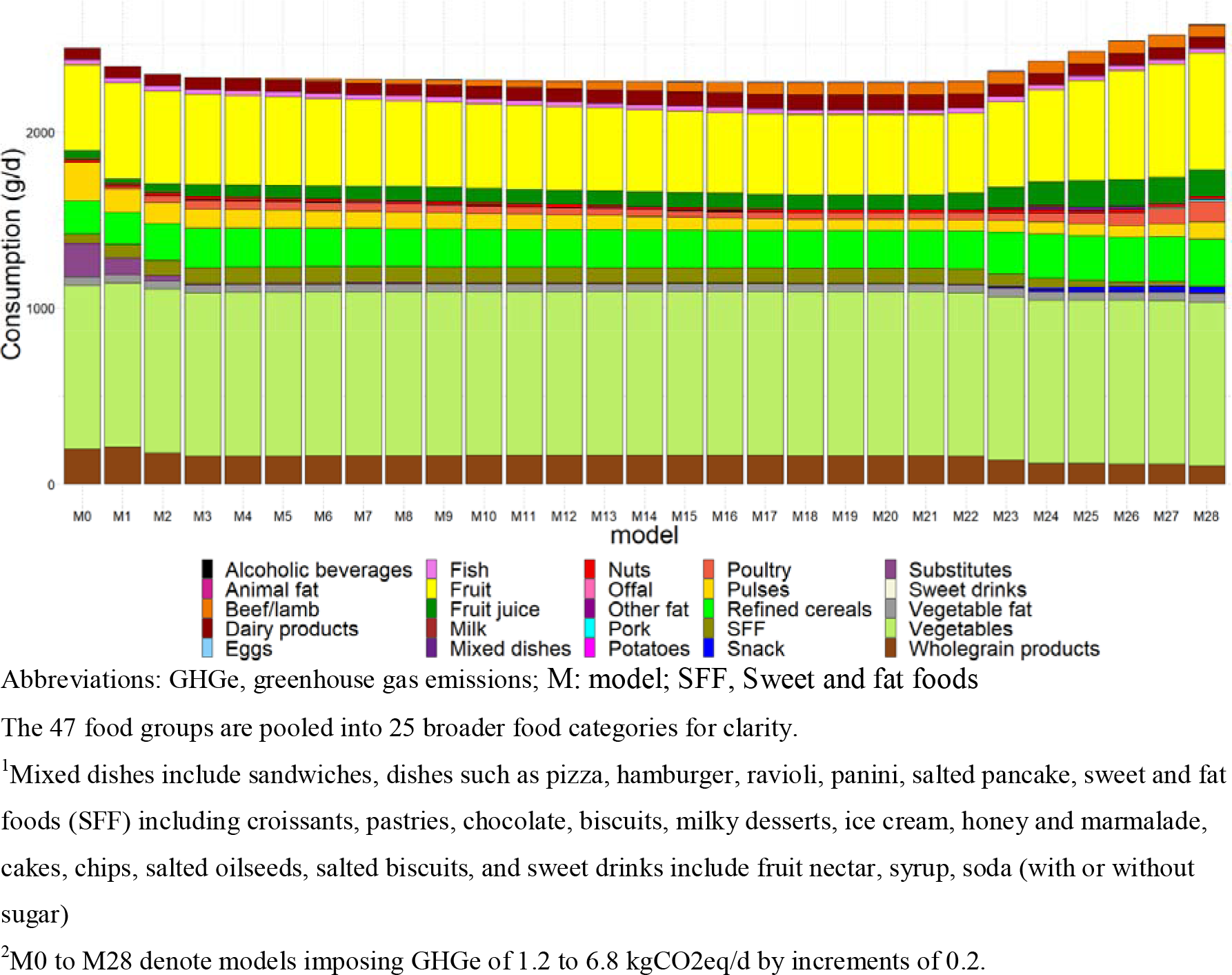
Food group consumptions (g/d) in modeled diets adhering to dietary guidelines of graded GHGE values ^1,2^

The Figure 3 details the contribution of food groups to total GHGe for the GHGe-imposed diets. The gradual increase in GHGe translated into a higher increase in the contribution of meat (beef/lamb, poultry, pork) and dairy products, from ∼30% (in M0) up to ∼85% (in M28).

**Figure 3:**
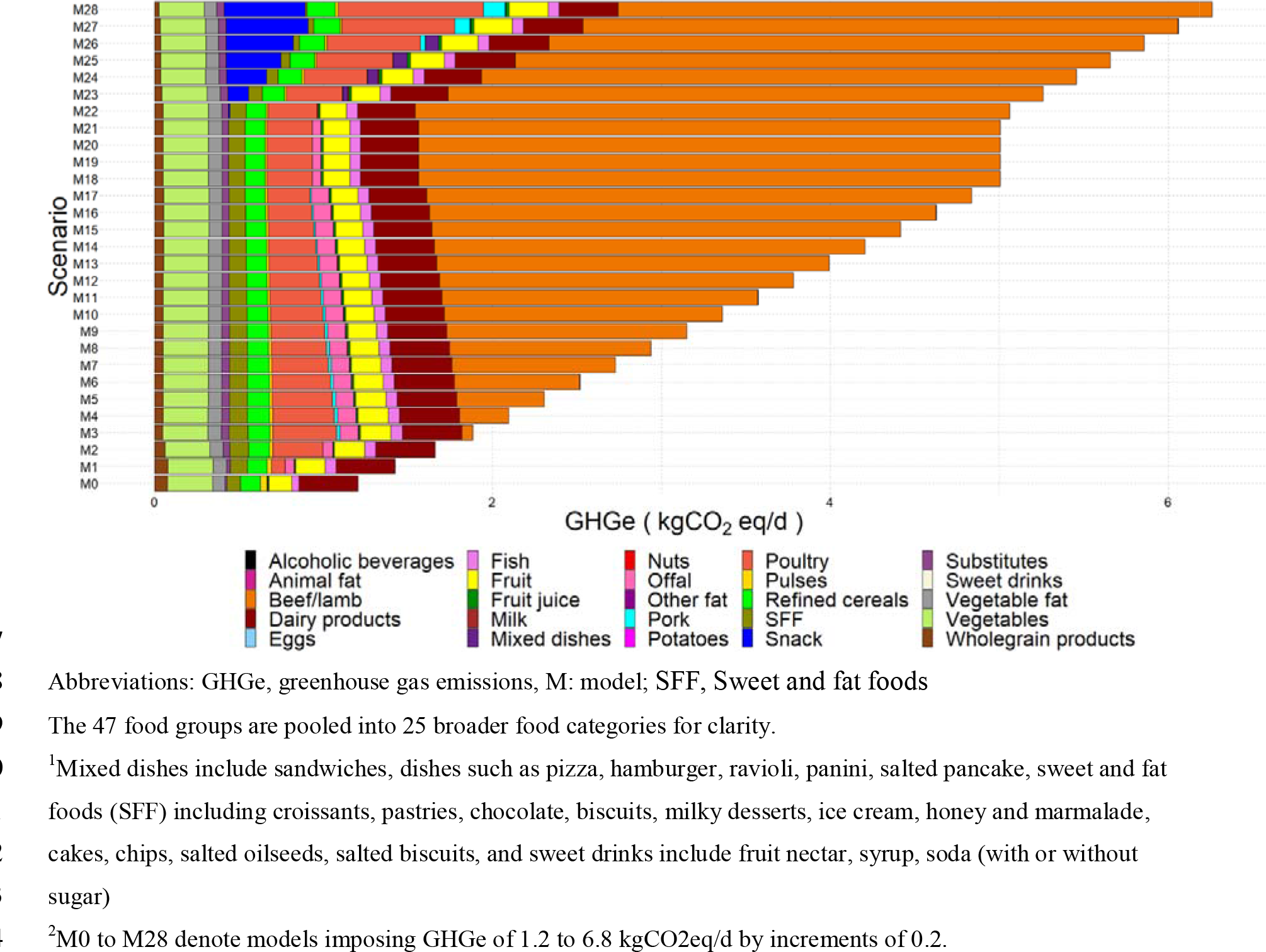
Contribution of food group to GHGe in GHG-imposed scenarios^1,2^

**Supplemental** Figure 1 shows, for illustrative purposes, the contributions of food groups to nutrient intakes across different modeled diets.

In the sensitivity analyses, decreasing the upper limit for total meat consumption from 500 to 200g/wk when identifying the healthy diets induced a decrease in their maximum total diet-related GHGe value, from 6.44 (M500) to 4.38 kgCO_2_eq/d (M200) **Table 2**). There were concomitant decreases in land occupation, energy demand, and HRS, while the percentage of plant protein and the percentage of organic food increased. The optimized diets were similar, except for a decrease in cereals, substitutes, and meat (regardless of type), and an increase in pulses and whole grains. To comply with nutritional references, beef/lamb was selected while poultry and pork were excluded. Additionally, lowering the maximum amount of food that could be consumed from the 99th to the 95th percentile in the acceptability constraints had only a slight impact on the results (**Supplemental Table 5**). The differences were minor, mostly affecting the diets with high GHGe. For example, the amount of vegetables decreased, and there was a shift towards more pulses, and the amount of poultry decreased, resulting in no solution beyond 5.6 kgCO2eq/d.

**Table 2:** Description of the diet models, maximizing GHGe constrained for different levels of total meat. ^1^

|  | M500 | M400 | M300 | M200 |
| --- | --- | --- | --- | --- |
| GHGe (kgCO<sub>2</sub>eq/d) | 6.44 | 5.75 | 5.07 | 4.38 |
| Land occupation (m<sup>2</sup>/d) | 18.47 | 16.25 | 14.02 | 11.81 |
| Cumulative energy demand (MJ/d) | 29.15 | 27.81 | 26.44 | 25.06 |
| Energy intake (kcal/d) | 2433.69 | 2405.18 | 2375.69 | 2373.86 |
| % organic food in the diet | 24 | 24 | 24 | 27 |
| HRS<sup>2</sup> | 0.39 | 0.34 | 0.30 | 0.20 |
| Plant protein (% of total protein) | 53.27 | 54.96 | 56.80 | 60.16 |
| **Consumption (g/d)** |  |  |  |  |
| Alcoholic beverages | 1 | 1 | 1 | 1 |
| Animal fat | 0 | 0 | 0 | 0 |
| Beef | 71 | 57 | 43 | 29 |
| Cereals | 306 | 295 | 282 | 247 |
| Dairy products | 102 | 102 | 102 | 102 |
| Eggs | 0 | 0 | 0 | 0 |
| Fish | 29 | 29 | 29 | 29 |
| Fruit | 637 | 637 | 637 | 634 |
| Fruit juice | 150 | 150 | 150 | 150 |
| Milk | 0 | 0 | 0 | 0 |
| Nuts | 15 | 15 | 15 | 15 |
| Offal | 0 | 0 | 0 | 0 |
| Mixed dishes<sup>3</sup> | 76 | 82 | 88 | 92 |
| Other fat | 0 | 0 | 0 | 0 |
| Pork | 0 | 0 | 0 | 0 |
| Potatoes | 0 | 0 | 0 | 0 |
| Poultry | 0 | 0 | 0 | 0 |
| Pulses | 57 | 57 | 57 | 99 |
| SFF<sup>4</sup> | 0 | 0 | 0 | 0 |
| Snack | 49 | 49 | 49 | 49 |
| Sweet drinks<sup>5</sup> | 0 | 0 | 0 | 0 |
| Substitutes | 41 | 37 | 33 | 28 |
| Vegetable fat | 45 | 44 | 44 | 44 |
| Vegetables | 930 | 930 | 930 | 930 |
| Wholegrain products | 215 | 226 | 239 | 274 |

Abbreviations: GHGe, greenhouse gas emissions; HRS, health risk score; SFF, Sweet and fat foods
<sup>1</sup> All models maximized GHGe under nutritional, dietary guidelines, and acceptability constraints. M500 to M200 referred to a maximum of 500 to 200 g of meat per week
<sup>2</sup>HRS (%) is the normalized distance to the theoretical minimum-risk exposure levels (TMREL) from the Global Burden of Diseases, expressed in % (i.e., HRS = 0% when the diet is at minimal risk by meeting all the TMREL and HRS=100% when the diet is at maximal risk by deviating from them at most)
<sup>3</sup>Mixed dishes include sandwiches, dishes such as pizza, hamburger, ravioli, panini, salted pancake
<sup>4</sup>Sweet and fat foods (SFF) including croissants, pastries, chocolate, biscuits, milky desserts, ice cream, honey and marmalade, cakes, chips, salted oilseeds, salted biscuits
<sup>5</sup>Sweet drinks include fruit nectar, syrup, soda (with or without sugar)

## Discussion

### GHG of French FBDG as compared to others

In the present study, we observed that it was possible to obtain diets nutritionally adequate and adhering to all recommandations of the French FBDG with associated GHGe ranging from 1.6 to 6.8 kgCO^2^eq/d. This large GHGe range can be explained by the fact that the French FBDG do not have a low specific target for total meat but only recommends upper limits for red and processed meats that are relatively high (e.g. 500g/wk) compared to other FBDGs, especially in countries where a strong emphasis already exists to promote environmental sustainability alongside health. For instance, in the Netherlands, it is recommended that individuals limit their consumption of all types of meat (i.e. including poultry) to 500 grams per week ^12^. Although the production of meat from poultry generates less GHG than ruminant meat, their emissions per kilogram are still significant and much higher than those of plant-based foods ^3,6,40^.

Our results are consistent with the large body of literature showing that consumption of animal products, especially meat is associated with very high GHGe ^41–43^. This is the case even for diets following FBDG. For example, a study conducted in the Netherlands, based on the recommendations before they were updated, found that following dietary guidelines could reduce the environmental impact of males aged 31-50y by up to 13%, while it might increase it by up to 5% for women aged 19- 30y. Conversely, adopting a meat-free version of the same diet according to the Dutch guidelines could reduce the environmental impact by 28% to 46% ^44^. Likewise, following the Dietary Guidelines for Americans for an omnivore diet does not necessarily result in lower GHGe, mostly because of the high levels of total meat, in sharp contrast to the vegetarian version of the Dietary Guidelines for Americans ^45^.

Our results, along with others, underscore that following the French FBDG can result in a large range of environmental pressures. For this eason, some countries have directly (i.e. included in diet modeling) considered the environmental criteria when assessing FBDG ^11^, particularly GHGe, when developing their dietary guidelines, unlike France. For instance, recently the Netherlands has based its guidelines on optimization models that establish maximum consumption levels for foods that emit high levels of greenhouse gases ^12^. The United States has developed guidelines for broad food groups, such as the Protein Foods Group, which includes lean meat and poultry, eggs, seafood, beans, peas, and lentils, nuts, seeds, and soy-based products. This has resulted in vastly different environmental footprints for the set of diets that comply with the guidelines for the “Protein Foods Group” depending on the type of food in that group ^46^.

### Levers of the FBDG on GHG and healthiness

Here, we found that complying with FBDG while departing as less as possible from the usual diet led to a ∼4%/1000Kcal GHGe increase compared to the observed diet. Thus, people wanting to increase their adherence to FBDG with a minimal effort from the current French diet may slightly increase climate pressure. This result is consistent with the extensive scientific literature reporting that all the healthy diets are not necessarily low-emission diets ^14,47,48^ and that there are large variations in GHGe across FBDGs ^14,49^.

When GHGe was also constrained, results showed that plant-based diets led to lower emissions than those containing more or less substantial amounts of animal products, especially ruminant meat, in line with the scientific literature ^41–43^. This is also consistent with recent work focusing on protein, showing that a healthy diet (in terms of both nutritional adequacy and long-term health) richer in plant protein led to lower environmental pressures ^50^. In addition, our long-term health indicator (reflecting adherence to the 2019 Global Burden of Diseases’s TMRELs) showed that, within the limits of the FBDGs, a more plant-based diet, rich in fruit and vegetables, pulses, and wholegrains, was associated with a lower health risk. This is in line with the literature documenting the health value of more plant- based diets ^40,51,52^. It also highlights the fact that diets following dietary recommendations exhibit a wide range of health risks.

### Other issues remaining unresolved and implications

GHGe is generally considered a good marker of global environmental pressures ^53^. However, the climate mitigation approach should not overlook other equally important indicators for achieving sustainable food systems, particularly water use, biodiversity conservation, and fisheries resources. Indeed, we recently showed in an analysis of the trade-offs between reducing water use and reducing GHGe that there are some discrepancies between modeled diets depending on whether the modeling is guided by one parameter or the other ^54^. Indeed, plant-based diets are generally better for both health and the environment, but there are still possible conflicts between certain environmental criteria, particularly water use ^55^. Then, as diets rich in plant-based food may increase some exposure to chemicals ^56^, such as synthetic pesticide residues, other factors, such as pollutants, could also be included in models to limit health risks ^16^. Finally, modeled healthy low-GHGe diets, which are rich in plant foods, were characterized by selecting organic foods in preference to conventional foods, as they exhibit lower GHGe, as previously documented ^57^. Thus, in the context of climate mitigation, it is important to consider not only dietary patterns but also production methods and possible improvements in agricultural practices. Also, optimized diets, which prioritize lower emissions and increased levels of plant products as organic, often come at a higher cost ^27^. Even though it would reduce their exposure to synthetic pesticides, this raises concerns about affordability for consumers.

### Assessment of FBDG in relation to the FAO principles

Beyond addressing environmental impacts, the FAO principles establish a list of targets to promote food sustainability ^18^. In that context, several studies have recently evaluated the sustainability of official FBDG across different countries ^11,14,19,20^. In the study conducted by James-Martin et al. ^11^, evaluating compliance with the 16 FAO principles for a sustainable healthy diet ^18^, France scored poorly because it did not numerically consider environmental criteria while setting their dietary guidelines and omitted other principles. On the other hand, the Belgian guidelines received the best score for the consumer official document.

In another report, a climate change score was assigned to the guidelines from 93 countries ^19^. Here again, Belgian dietary guidelines received the best score (84/100), while the French ones were rated lower (51/100). The latter score was mostly undermined by the absence of any reference to animal product substitution. Finally, the guidelines on animal products, and therefore the scope for consumption of these food groups, appear to be a key lever for ensuring the sustainability of appropriate diets, particularly in environmental terms.

### Strengths and Limitations

Our study has a few limitations. Because the people who participated in the study were all volunteers who were presumably more interested in nutritional matters, their initial diet before optimization was already quite rich in plant-based foods compared to what is typically observed in the general population. This has probably led to higher 99^th^ percentile values than in a representative sample. The LCA only considered the production stage as data for the entire system (from farm to fork) was not accessible for organic systems. However, whether for organic or standard/conventional farming systems for LCA, which has rarely been considered before, the production phase has the highest emissions ^58^.

Additionally, it has been established that the LCA may inaccurately represent some ecosystem services, particularly for agroecological practices ^59^. It would be also valuable to consider other environmental indicators, as discussed above, and consequential LCA. Here, the consequences in terms of reshaping agricultural practices and mitigation associated with lower production of animal products are not considered.

Our study has many strengths. When modeling diets, we considered many nutrient reference values, including bioavailability for iron and zinc and cultural “acceptability,” which corresponds to the apparent feasibility of the solutions. We also used recent and reliable data from the GBD as a proxy for the potential impact of the diet on health.

## Conclusion

In conclusion, this study highlights specific dietary adjustments that can significantly reduce the environmental footprint of diets while also providing health benefits. According to scientific literature, dietary changes alone could reduce environmental impact by up to 80%. A key adjustment involves redefining the role of meat in dietary guidelines, including the introduction of thresholds for different types of meat, with a particular focus on ruminant meat. To achieve truly sustainable diets, a multidisciplinary approach is essential. This approach should consider a range of factors beyond greenhouse gas emissions, addressing various environmental, health, and socio-economic issues.

## Data Availability

Data described in the manuscript, code book, and analytic code will be made available upon request pending application and approval. Researchers from public institutions can submit a collaboration request including information on the institution and a brief description of the project to. All requests will be reviewed by the steering committee of the NutriNet-Sante study. If the collaboration is accepted, a data access agreement will be necessary and appropriate authorizations from the competent administrative authorities may be needed. In accordance with existing regulations, no personal data will be accessible.

## Acknowledgements

We thank Cédric Agaesse, Alexandre De Sa, Rebecca Lutchia (dietitians); Selim Aloui (IT manager), Thi Hong Van Duong, Selim Aloui (IT manager), Régis Gatibelza, Jagatjit Mohinder and Aladi Timera (computer scientists); Julien Allegre, Nathalie Arnault, Laurent Bourhis, Nicolas Dechamp, and Fabien Szabo de Edelenyi, PhD (supervisor) (data-manager/statisticians), Maria Gomes, Mirette Foham (participants’ support), Paola Yvroud, MD (health event validator), Marine Ricau (operational coordination), Nadia Khemache (HR and finance manager), Marie Ajanohun, Souad Hadji (administrative support) for their technical contribution to the NutriNet-Santé study and Marine Ricau (operational coordination). We warmly thank all the volunteers of the NutriNet-Santé cohort.

## The authors’ contributions are as follows

EKG, MT, and SH conducted the cohort study. EKG, SH, DL, PP and JB implemented databases. EKG, JB, HF, and FM, conducted the research.

EKG performed statistical analyses and drafted the manuscript.

All authors critically helped interpret results, revised the manuscript and provided relevant intellectual input. They all read and approved the final manuscript.

EKG had primary responsibility for the final content; she is the guarantor.

## Transparency statement

Dr Kesse-Guyot (the guarantor) affirms that the manuscript is an honest, accurate, and transparent account of the study being reported, that no important aspects of the study have been omitted and that any discrepancies from the study as planned have been explained.

## Code availability statement

Data described in the manuscript, code book, and analytic code will be made available upon request pending application and approval. Researchers from public institutions can submit a collaboration request including information on the institution and a brief description of the project to. All requests will be reviewed by the steering committee of the NutriNet-Santé study. If the collaboration is accepted, a data access agreement will be necessary and appropriate authorizations from the competent administrative authorities may be needed. In accordance with existing regulations, no personal data will be accessible.

