## Supplemental materials for "To be climate-friendly, food-based dietary guidelines must include limits on total meat consumption – modeling from the case of France"

Equipe de Recherche en Epidémiologie Nutritionnelle (EREN)

SMBH Université Sorbonne Paris Nord, 74 rue Marcel Cachin, 93017 Bobigny, France

### Supplemental Method 1: PNNS-GS2 computation

In March 2017, as part as the developmentof the fourth Programme National Nutrition Santé (2017-2021), the Haut Conseil de Santé Publique (HCSP) published a report updating the 2001 PNNS recommendations (5) based on scientific literature about the relationships between diet and long-term health and a model created by the Agence nationale de sécurité sanitaire de l'alimentation, de l'environnement et du travail (6). These new recommendations provide dietary guidelines, with 6 adequacy guidelines ("fruits and vegetables", "nuts", "legumes", "whole foods", "milk and dairy products", and "fish and seafood") and 6 moderation guidelines ("meat", "processed meat", "added fats", "sugary products", "beverages", and "salt"). Physical activity recommendations are not addressed by the HCSP. The nutrition experts who were involved in developing the guidelines defined the thresholds and corresponding scores. These thresholds and related scores are defined so asthat following the guidelines is associated with one point, whereas not following them is scored zero points for healthy foodss. To increase the power of discrimination, half-points are allocated in a linear fashion above the guideline thresholds. However, an exception was made for milk and dairy products, and fish. As, the relationship between these foods and health is non-linearpoints allocated points form a parabolic relationship (7).

The PNNS-GS2 emphasizes on the distinction between malus components (unhealthy food thought to be avoided, which have a negative moderation score, e.g. salt) and bonus components (healthy foods considered beneficial, which have a positive adequacy score, e.g. legumes).

All details regarding the scoring system are shown in **Supplemental Table 1.**

### Supplemental Method 2:Iron bioavailability

In order to consider iron bioavailability, heme and non-heme iron were considered (1).

The rate of absorption for heme iron was calculated as (2) :

$$\text{Log Absorption }\left( \text{\%} \right)\text{= 1.9897 – 0.3092 × log (SF) }$$

where SF is serum ferritin (μg/L). We considered a stringent situation by setting serum ferritin at 15 mg/L

The rate of absorption for non-heme iron was calculated as (3) :

$\text{Ln Absorption }\left( \text{\%} \right)\text{= }\text{6.294 – 0.709 }\text{ln}\text{ }\left( \text{SF} \right)\text{+ 0.119 ln }\left( \text{VitC} \right)\text{+ 0.006 ln }\left( \text{MFP + 0.1} \right)\text{ } \text{-0.055}\ln\left( \text{T+0.1} \right) \text{-0.247}\ln\left( \text{Phy} \right)\text{-0.137}\ln\left( \text{Ca} \right)\text{-0.083 ln (NHI)}$

where SF is serum ferritin (μg/L) which was also set at 15 mg/L, VitC is vitamin C intake (mg), MFP corresponds to consumption of meat, fish, and poultry (g), T is tea intake (as number of cups), Phy is phytate intake (mg), Ca is calcium intake (mg), and NHI is non-heme iron intake (mg).

### Supplemental Method 3: Zinc bioavailability

For zinc absorption, we used the equation developed and updated by Miller and al. (4)

$$BZ (mmol/d)=0.5 \left\{ 0.069 \left( 1+\frac{PhyI (1-0.017CI)}{0.44} \right)+ 0.084 + ZI (1+ 0.012PI) -\sqrt{\left( \left( 0.069(1+\frac{PhyI(1-0.017 CI)}{0.44}) + 0.084 + ZI (1+0.012PI \right)^{2}- 4\times0.084 ZI (1+0.012PI \right)} \right\}$$

Where BZ is bioavailable zinc and PhyI, CI, ZI, and PI are phytates, calcium, zinc and proteins daily intakes respectively. All variables are in units of mmol/d except protein which is in g/d.

### Supplemental Method 4: Health Risk Score computation

The Global burdent of diseases (GBD) study by the IHME aimed to identify health challenges worldwide (8). It defines theoretical minimum-risk exposure levels (TMREL) for unhealthy (red meat, processed meat and sweetened beverages) and healthy (wholegrain products, fruits, vegetables, pulses, nuts and seeds, and milk) food groups. The levels aim to estimate recommended target intakes for each of these groups. The study provides the DALYs associated with excessive or insufficient consumption of these food groups. A Health Risk Score (HRS) evaluates the health risks associated with a given diet (9). HRS is minimal when the unhealthy food group intake is lower or equal to TMREL value and healthy food group intake is greater or equal to TMREL value. The distance to each food group target is weighted by the relative importance of reaching this target compared to others in terms of DALYs. HRS is maximal when the diet is at the maximal possible distance from each TMREL value.

The following formula is as follows:

$$\text{HRS}=100*\left[ \sum_{i=1}^{3} \left\{ \frac{Cons\text{ }\left( i \right)}{Mean\_pop (i)}\times\frac{DALYs\text{ }\left( i \right)}{DALYs\text{ }\left( all \right)} \right\} + \sum_{j=1}^{6} \left\{ max\left( \frac{TMREL (j) - Cons\text{ }\left( j \right)}{TMREL (j)}; 0 \right)\times\frac{DALYs\text{ }\left( j \right)}{DALYs\text{ }\left( all \right)} \right\} \right]$$

*Where:*

- i: unhealthy food groups to be limited (red meat, processed meat and sweetened beverages)
- j: healthy food groups to be promoted (wholegrain products, fruits, vegetables, legumes, nuts and seeds, milk)
- Cons: consumption of the item i or j
- Mean_pop (i): average consumption of the food group i (g/d) in the French population
- TMREL (j): TMREL value for the food group j (g/d)
- DALYs (i), DALYs(j): DALYs associated with over- and under-consumptions of the food groups i and j respectively (years)
- DALYs (all): sum of all DALYs(i) and DALYs(j)

The TMREL and corresponding DALYs values used were the following:

|  |  | TMREL^1^ (g/d) | DALYs^2^ (y) |
| --- | --- | --- | --- |
| Healthy foods | Wholegrain products | 150 | 42 392 |
|  | Fruits | 325 | 29 643 |
|  | Legumes | 95 | 20 489 |
|  | Vegetables | 300 | 12 432 |
|  | Nuts and seeds | 14.5 | 7 885 |
|  | Milk | 430 | 6 248 |
| Unhealthy foods | Red meat | 0 | 49 386 |
|  | Processed meat | 0 | 20 634 |
|  | Sweetened beverages | 0 | 5 896 |
| Total |  |  | 195 006 |

^1^According to the most recent (2019) estimates from the GBD, TMREL values are global estimates corresponding to a mean energy intake of 2,300 kcal (8).

^2^ DALYs values associated with excessive/insufficient consumptions of unhealthy/healthy foods are available at the Global Health Data Exchange website (<http://ghdx.healthdata.org/gbd-results-tool>).

**Supplemental Table 1: Components, scoring and weighting used for sPNNS-GS2 computation**

| **Dietary components** | **Recommendation** | **Criteria** | **Score** |
| --- | --- | --- | --- |
| Fruit and | At least 5 serv/d, | [0 - 3.5[ | 0 |
| vegetables | with 1 max as juice and 1 | [3.5 - 5[ | 0.5 |
| (weight=3) | max as dried | [5 - 7.5[ | 1 |
|  |  | ≥7.5 | 2 |
| Nuts | A handful/d | 0 | 0 |
| (weight=1) |  | ]0 – 0.5[ | 0.5 |
|  |  | [0.5- 1.5[ | 1 |
|  |  | ≥1.5 | 0 |
| Pulses | At least 2 serv/w | 0 /w | 0 |
| (weight=1) |  | ]0-2[ /w | 0.5 |
|  |  | ≥2 /w | 1 |
| Whole-grain | Every day | 0 | 0 |
| Food |  | ]0 - 1[ | 0.5 |
| (weight=2) |  | [1 - 2[ | 1 |
|  |  | ≥2 | 1.5 |
| Milk and | 2 serv/d | [0 - 0.5[ | 0 |
| dairy products |  | [0.5 - 1.5[ | 0.5 |
| (weight=1) |  | [1.5 - 2.5[ | 1 |
|  |  | ≥2.5 | 0 |
| Red meat | Limit consumption | ≥750 g/w | -2 |
| (weight=2) |  | [500 - 750[ g/w | -1 |
|  |  | [0 - 500[ g/w | 0 |
| Processed meat | Limit consumption | ≥300 g/w | -2 |
| (weight=3) |  | [150 - 300[ g/w | -1 |
|  |  | [0 - 150[ g/w | 0 |
| Fish and | 2 serv/w | [0 - 1.5[serv /w | 0 |
| Seafood |  | [1.5 - 2.5[serv /w | 1 |
| (weight=2) |  | [2.5 - 3.5[serv /w | 0.5 |
|  |  | ≥3.5 serv /w | 0 |
| Added fat | Avoid overeating | >16% of EIWA ^c^ | 0 |
| (weight=2) |  | ≤16% of EIWA | 1.5 |
| Sugary foods | Limit consumption | ≥15% of EIWA | -2 |
| (weight=3) |  | [10-15[% of EIWA | -1 |
|  |  | <10 % of EIWA | 0 |
| Sweet drinks | Limit consumption | ≥ 750mL mL/d | -2 |
| Beverages |  | [250 - 750[ mL/d | -1 |
| (weight=3) |  | ]0 - 250[ mL/d | -0.5 |
|  |  | 0 mL/d | 0 |
| Alcoholic | Limit consumption | >200 g/d | -2 |
| beverages |  | ]100-200] g/d | -1 |
| (weight=3) |  | ]0-100] g/d | 0 |
|  |  | 0 g/d | 0.5 |
| Salt | Limit consumption | >12 g/d | -2 |
| (weight=3) |  | ]10-12] g/d | -1 |
|  |  | ]8-10] g/d | -0.5 |
|  |  | ]6-8] g/d | 0 |
|  |  | ≤6 g/d | 1 |

Abbreviations: d: day; EIWA: Energy intake without alcohol; w: week;

### Supplemental Table 2: Nutritional constraints used in the optimization models

|  | **Men** |  | **Women** |  | **Average individual^1^** |  |
| --- | --- | --- | --- | --- | --- | --- |
|  | **Lower reference** | **Upper reference** | **Lower reference** | **Upper reference** | **Lower reference** | **Upper reference** |
| EI | ER - 8% | ER + 8% | ER - 8% | ER + 8% | ER - 8% | ER + 8% |
| Protein | 0.83 × bw g | 2.3 × bw g | 0.83 × bw g | 2.3 × bw g | 0.83 × bw g | 2.3 × bw g |
| Vitamin A | 750 µg | 3000 µg | 650 µg | 3000 µg | 700 µg | 3000 µg |
| Vitamin B1 | 0.42 µg /kcal | - | 0.42µg /kcal | - | 0.42µg /kcal | - |
| Vitamin B2 | 1.60 mg | - | 1.60 mg | - | 1.60 mg | - |
| Vitamin B3 | 6.7 µg /kcal | 900 µg | 6.7 µg /kcal | 900 µg | 6.7 µg /kcal | 900 µg |
| Vitamin B5 | P5 mg | - | P5 mg | - | Weighted P5 | - |
| Vitamin B6 | 1.7 mg | 25 mg | 1.6 mg | 25 mg | 1.65 mg | 25 mg |
| Vitamin B9 | 330 µg | - | 330 µg | - | 330 µg | - |
| Vitamin B12 | 4 µg | - | 4 µg | - | 4 µg | - |
| Vitamin C | 110 mg | - | 110 mg | - | 110 mg | - |
| Vitamin E | P5 g | - | P5 g | - | Weighted P5 | - |
| Vitamin K | P5 µg | - | P5 µg | - | Weighted P5 | - |
| Calcium | 950 mg | 2500 mg | 950 mg | 2500 mg | 950 mg | 2500 mg |
| Copper | P5 g | 5 g | P5 g | 5 g | Weighted P5 | 5 g |
| Bioavailable Iron^2^ | 1.10 mg | - | 1.10/1.16 mg | - | 1.13 mg |  |
| Iodine | 150 µg | 600 µg | 150 µg | 600 µg | 150 µg | 600 µg |
| Magnesium | P5 g | - | P5 g | - | Weighted P5 | - |
| Manganese | P5 g | - | P5 g | - | Weighted P5 | - |
| Phosphorus | 550 mg | - | 550 mg | - | 550 mg | - |
| Potassium | 3500 mg | - | 3500 mg | - | 3500 mg | - |
| Selenium | 70 µg | 300 µg | 70 µg | 300 µg | 70 µg | 300 µg |
| Sodium | 1500 mg | 2300 mg | 1500 mg | 2300 mg | 1500 mg | 2300 mg |
| Bioavailable zinc | 1.6 mg |  | 1.3 mg |  | 1.45 mg |  |
| SFA | - | 12% EI | - | 12% EI | - | 12% EI |
| Linoleic acid | 4% EI | - | 4% EI | - | 4% EI | - |
| ALA | 1% EI | - | 1% EI | - | 1% EI | - |
| LA / ALA | - | 5 | - | 5 | - | 5 |
| EPA+DHA | 0.5 g | - | 0.5 g | - | 0.5 g | - |
| Sugar without lactose | - | 100 g | - | 100 g | - | 100 g |
| Fiber | 30 g | - | 30 g | - | 30 g | - |

Abbreviations: ALA, alpha-linoleic acid; bw, body weight (kg); DHA, docosahexaenoic acid; EI, energy intake; EPA, eicosapentaenoic acid; ER, energy requirement; LA, linolenic acid; M^-^, non-menopausal; M^+^, menopausal; SFA, saturated fatty acids

^1^In case of different references between men and women. The average individual was the weighted mean as follows: 50% men, 25% women M-, 25% M+.

^2^threshold corresponding to a deficiency prevalence ≤5%

^3^threshold corresponding to the deficiency cut-off

### Supplemental Table 3: Characteristics of the Sample (N=29,413) by weighted quantiles of PNNS-GS2 scores^1^

|  | Q1 | Q2 | Q3 | Q4 | Q5 | Ptrend^2^ |
| --- | --- | --- | --- | --- | --- | --- |
| **sPNNS-GS2** | -2.72 (2.24) | 0.52 (1.25) | 2.38 (1.06) | 4.23 (0.94) | 6.90 (1.34) | <0.001 |
| **Gender (%)** |  |  |  |  |  |  |
| Women | 50.2 | 50.4 | 49.8 | 49.8 | 50.2 | 0.83 |
| Men | 49.8 | 49.6 | 50.2 | 50.2 | 49.8 |  |
| **Age** | 54.26 (13.78) | 54.58 (14.09) | 54.68 (14.15) | 55.39 (13.85) | 56.02 (13.66) | <0.001 |
| **Education (%)** |  |  |  |  |  |  |
| < High-school diploma | 59.8 | 63.3 | 64.2 | 64.5 | 64.8 | <0.001 |
| High school diploma | 14.6 | 14.1 | 13.8 | 13.4 | 13.4 |  |
| Postgraduate | 25.6 | 22.6 | 22 | 22.1 | 21.7 |  |
| **Income^3^ (%)** |  |  |  |  |  |  |
| <900 € | 6.9 | 6.2 | 5.5 | 5.8 | 6.7 |  |
| 900 – 1,200 € | 5.6 | 4.3 | 4 | 3.6 | 4 |  |
| 1,200 – 1,800 € | 24.6 | 24 | 22.5 | 21.5 | 19.9 | <0.001 |
| 1,800 – 2,300 € | 15.6 | 15.2 | 16.2 | 16.4 | 15.6 |  |
| 2,300 – 2,700 € | 11.4 | 10.8 | 12.2 | 12 | 11.5 |  |
| 2,700 – 3,700 € | 19.1 | 20.1 | 19.3 | 20.3 | 20.2 |  |
| ≥3700 € | 12.2 | 14.2 | 15.6 | 15.2 | 16.5 |  |
| **Occupation status (%)** |  |  |  |  |  |  |
| Unemployed | 3.8 | 3.8 | 3.6 | 3.7 | 3.7 | 0.66 |
| Retired | 40.1 | 41.3 | 41.9 | 43 | 43.5 |  |
| Employees | 15.1 | 12.9 | 12 | 10.8 | 10.1 |  |
| Intermediate professions | 13 | 13.4 | 13.8 | 13.6 | 12.3 |  |
| Manger or intellectual | 20.1 | 20.9 | 22.3 | 22.3 | 23.1 |  |
| never employed | 5.9 | 5.7 | 4.7 | 4.8 | 5.4 |  |
| self-employed | 2.1 | 2 | 1.7 | 1.7 | 1.9 |  |
| **Marital status (%)** |  |  |  |  |  |  |
| Cohabiting | 90 | 90.2 | 88.3 | 87.7 | 86 | <0.001 |
| Single | 10 | 9.8 | 11.7 | 12.3 | 14 |  |
| **Smoking status (%)** |  |  |  |  |  |  |
| Former | 45.6 | 44.6 | 42.9 | 43.9 | 43.9 | <0.001 |
| Current | 14.8 | 10.8 | 10.6 | 9.2 | 6.4 |  |
| Never | 39.7 | 44.7 | 46.4 | 46.9 | 49.7 |  |
| **Physical activity (%)** |  |  |  |  |  |  |
| High | 34.9 | 34.2 | 34.6 | 37.3 | 39.7 | <0.001 |
| Medium | 32.7 | 34.4 | 36 | 35.5 | 36.9 |  |
| Low | 22.3 | 21 | 19.2 | 16.9 | 13.3 |  |
| **Body mass index (kg/m²)** | 25.64 (4.80) | 24.89 (4.55) | 24.45 (4.40) | 24.24 (4.23) | 23.57 (4.02) | <0.001 |
| **Energy intake (Kcal/d)** | 2614 (699) | 2214 (618) | 2009 (576) | 1849 (534) | 1725 (460) | <0.001 |
| **%organic food** | 0.19 (0.20) | 0.23 (0.23) | 0.27 (0.26) | 0.32 (0.28) | 0.41 (0.31) | <0.001 |
| **% plant protein** | 24.88 (7.86) | 28.69 (9.71) | 32.34 (12.04) | 36.28 (14.62) | 43.51 (17.91) | <0.001 |
| **GHGe(kgCO_2_eq/d)/1000 Kcal** | 2.54 (0.97) | 2.26 (0.96) | 2.07 (0.90) | 1.86 (0.85) | 1.60 (0.76) | <0.001 |
| **LO(m²)** | 17.10 (9.40) | 12.91 (7.36) | 10.71 (5.86) | 8.92 (4.75) | 7.25 (3.58) | <0.001 |
| **CED (MJ/d)** | 25.48 (9.04) | 20.32 (7.52) | 17.51 (6.33) | 15.44 (5.70) | 13.61 (4.68) | <0.001 |
| **GHGe (kgCO_2_eq/d)** | 6.64 (3.33) | 4.98 (2.63) | 4.09 (2.08) | 3.37 (1.73) | 2.68 (1.32) | <0.001 |
| **Consumption (g/d)** |  |  |  |  |  |  |
| Alcoholic beverages | 219.69 (241.12) | 142.64 (170.88) | 119.69 (167.92) | 98.46 (160.71) | 62.92 (80.65) | <0.001 |
| Animal fat | 8.64 (8.63) | 6.83 (7.29) | 5.84 (6.18) | 5.21 (5.95) | 4.08 (5.09) | <0.001 |
| Beef | 73.81 (54.87) | 51.24 (43.02) | 40.22 (35.67) | 30.89 (30.08) | 21.82 (21.49) | <0.001 |
| Cereals | 178.34 (102.61) | 154.71 (99.67) | 139.65 (93.47) | 124.68 (93.85) | 105.36 (86.24) | <0.001 |
| Dairy products | 216.11 (154.38) | 199.94 (141.44) | 183.73 (135.30) | 170.12 (129.88) | 153.20 (125.64) | <0.001 |
| Eggs | 13.40 (13.66) | 11.63 (12.13) | 11.05 (12.16) | 10.37 (11.16) | 10.38 (11.91) | <0.001 |
| Fish | 57.35 (59.62) | 49.07 (42.76) | 46.47 (41.08) | 45.48 (43.33) | 40.35 (37.74) | <0.001 |
| Fruit | 227.59 (210.74) | 260.21 (252.42) | 267.60 (249.74) | 289.00 (244.69) | 367.33 (273.71) | <0.001 |
| Fruit juice | 100.62 (137.41) | 91.07 (115.56) | 88.91 (118.76) | 83.60 (112.83) | 73.82 (101.25) | <0.001 |
| Milk | 72.65 (147.88) | 68.13 (148.61) | 62.97 (134.33) | 53.23 (128.84) | 39.74 (111.29) | <0.001 |
| Nuts | 4.88 (11.17) | 5.92 (14.21) | 7.81 (17.35) | 8.76 (16.80) | 13.25 (19.69) | <0.001 |
| Offal | 3.12 (5.87) | 2.49 (9.16) | 1.90 (10.72) | 1.65 (3.88) | 1.25 (3.02) | <0.001 |
| Mixed dishes^4^ | 41.02 (62.32) | 33.79 (34.10) | 27.34 (22.40) | 23.92 (20.64) | 20.28 (19.69) | <0.001 |
| Other fat | 10.30 (10.07) | 8.24 (8.96) | 7.03 (8.22) | 6.02 (6.95) | 5.00 (6.73) | <0.001 |
| Pork | 95.55 (60.48) | 63.61 (40.49) | 45.25 (28.67) | 32.37 (21.87) | 20.95 (17.20) | <0.001 |
| Potatoes | 33.99 (29.43) | 27.00 (24.87) | 23.44 (25.30) | 20.57 (20.63) | 17.37 (17.30) | <0.001 |
| Poultry | 34.37 (30.76) | 26.99 (24.76) | 23.18 (22.75) | 20.29 (23.06) | 17.52 (23.14) | <0.001 |
| Pulses | 15.30 (23.61) | 15.37 (33.42) | 15.37 (25.32) | 16.96 (29.81) | 23.38 (41.88) | <0.001 |
| SFF^5^ | 94.96 (76.69) | 81.62 (60.03) | 71.93 (51.55) | 63.46 (44.72) | 53.63 (37.82) | <0.001 |
| Snack | 17.88 (26.43) | 12.07 (13.23) | 10.31 (12.91) | 8.05 (10.29) | 5.79 (8.53) | <0.001 |
| Sweet drinks^6^ | 85.55 (167.96) | 52.44 (122.41) | 40.44 (86.21) | 32.98 (82.91) | 21.82 (49.45) | <0.001 |
| Substitutes | 17.43 (86.41) | 22.57 (107.63) | 35.14 (133.74) | 47.21 (155.87) | 76.12 (176.15) | <0.001 |
| Vegetable fat | 27.20 (18.96) | 23.65 (17.00) | 22.00 (15.67) | 20.85 (15.14) | 18.99 (13.73) | <0.001 |
| Vegetables | 338.92 (225.09) | 344.02 (231.34) | 342.90 (253.72) | 348.97 (221.61) | 397.51 (243.20) | <0.001 |
| Wholegrain products | 38.81 (60.33) | 49.03 (72.69) | 59.12 (77.11) | 65.47 (78.41) | 78.43 (77.20) | <0.001 |

Abbreviations: CED Cumulative energy demand; GHGe, greenhouse gas emissions; LO, land use; SFF, Sweet and fat foods

^1^Values presented are means (SD) and are weighted on the individual weight to represent the average individual.

^2^Chi² test or Kruskal-Wallis test as appropriate

^3^Income per consumption unit per month

^4^Mixed dishes include sandwiches, dishes such as pizza, hamburger, ravioli, panini, salted pancake

^5^Sweet and fat foods (SFF) include croissants, pastries, chocolate, biscuits, milky desserts, ice cream, honey and marmalade, cakes, chips, salted oilseeds, salted biscuits

^6^Sweet drinks include fruit nectar, syrup, soda (with or without sugar)

### Supplemental Table 4: Consumption of food groups in GHG-imposed scenarios^1^

|  |  | M0 | M1 | M2 | M3 | M4 | M5 | M6 | M7 | M8 | M9 | M10 | M11 | M12 | M13 | M14 | M15 | M16 | M17 | M18 | M19 | M20 | M21 | M22 | M23 | M24 | M25 | M26 | M27 | M28 |
| --- | --- | --- | --- | --- | --- | --- | --- | --- | --- | --- | --- | --- | --- | --- | --- | --- | --- | --- | --- | --- | --- | --- | --- | --- | --- | --- | --- | --- | --- | --- |
| **Alcoholic beverages** | alcoholic beverages | 1 | 1 | 1 | 1 | 1 | 1 | 1 | 1 | 1 | 1 | 1 | 1 | 1 | 1 | 1 | 1 | 1 | 1 | 1 | 1 | 1 | 1 | 1 | 1 | 1 | 1 | 1 | 1 | 1 |
| **Animal fat** | butter, lard etc. | 0 | 0 | 0 | 0 | 0 | 0 | 0 | 0 | 0 | 0 | 0 | 0 | 0 | 0 | 0 | 0 | 0 | 0 | 0 | 0 | 0 | 0 | 0 | 0 | 0 | 0 | 0 | 0 | 0 |
| **Beef** | beef/lamb, veal | 0 | 0 | 1 | 6 | 10 | 15 | 20 | 24 | 29 | 33 | 38 | 43 | 47 | 52 | 56 | 61 | 65 | 70 | 70 | 70 | 70 | 71 | 71 | 71 | 71 | 71 | 71 | 71 | 72 |
| **Refined cereals** | breakfast cereals | 27 | 18 | 12 | 12 | 12 | 11 | 11 | 10 | 10 | 9 | 9 | 8 | 8 | 7 | 7 | 6 | 6 | 5 | 5 | 5 | 5 | 4 | 2 | 0 | 0 | 0 | 0 | 0 | 0 |
|  | refined bread | 0 | 22 | 33 | 33 | 32 | 32 | 32 | 32 | 33 | 34 | 35 | 36 | 36 | 37 | 38 | 39 | 40 | 42 | 42 | 42 | 42 | 45 | 46 | 46 | 36 | 26 | 16 | 0 | 0 |
|  | refined cereal | 155 | 169 | 179 | 178 | 177 | 176 | 176 | 174 | 173 | 172 | 171 | 170 | 169 | 168 | 167 | 166 | 165 | 165 | 165 | 165 | 165 | 167 | 187 | 206 | 217 | 227 | 239 | 265 | 248 |
| **Dairy products** | cheese | 63 | 62 | 62 | 62 | 62 | 62 | 62 | 61 | 61 | 60 | 60 | 59 | 59 | 58 | 58 | 57 | 57 | 56 | 56 | 56 | 56 | 56 | 58 | 59 | 61 | 62 | 62 | 63 | 63 |
|  | cottage cheese | 0 | 0 | 0 | 0 | 0 | 0 | 0 | 0 | 0 | 0 | 0 | 0 | 0 | 0 | 0 | 0 | 0 | 0 | 0 | 0 | 0 | 0 | 0 | 0 | 0 | 0 | 0 | 0 | 0 |
|  | petits suisses | 0 | 3 | 5 | 5 | 5 | 5 | 5 | 5 | 5 | 5 | 5 | 4 | 4 | 4 | 4 | 4 | 4 | 4 | 4 | 4 | 4 | 4 | 5 | 6 | 6 | 5 | 4 | 0 | 0 |
|  | yogurt | 0 | 0 | 0 | 0 | 0 | 0 | 0 | 0 | 3 | 6 | 8 | 11 | 13 | 16 | 18 | 20 | 22 | 24 | 24 | 24 | 24 | 22 | 9 | 0 | 0 | 0 | 0 | 0 | 0 |
| **Eggs** | eggs | 0 | 0 | 0 | 0 | 0 | 0 | 0 | 0 | 0 | 0 | 0 | 0 | 0 | 0 | 0 | 0 | 0 | 0 | 0 | 0 | 0 | 0 | 0 | 0 | 0 | 0 | 0 | 0 | 0 |
| **Fish** | crustacean | 2 | 1 | 1 | 1 | 1 | 1 | 1 | 1 | 1 | 1 | 1 | 1 | 1 | 1 | 1 | 1 | 1 | 1 | 1 | 1 | 1 | 1 | 1 | 1 | 1 | 1 | 1 | 1 | 0 |
|  | fat fish | 16 | 16 | 16 | 16 | 16 | 16 | 16 | 16 | 16 | 16 | 16 | 16 | 16 | 16 | 16 | 16 | 16 | 16 | 16 | 16 | 16 | 16 | 16 | 16 | 16 | 16 | 16 | 16 | 16 |
|  | other fish | 11 | 11 | 12 | 12 | 12 | 12 | 12 | 12 | 12 | 11 | 11 | 11 | 11 | 11 | 11 | 11 | 11 | 11 | 11 | 11 | 11 | 11 | 11 | 11 | 11 | 12 | 12 | 12 | 13 |
| **Fruit** | compote | 3 | 4 | 4 | 5 | 5 | 5 | 6 | 6 | 7 | 7 | 7 | 8 | 8 | 9 | 9 | 9 | 10 | 10 | 10 | 10 | 10 | 10 | 10 | 10 | 10 | 9 | 5 | 0 | 0 |
|  | dried fruits | 0 | 0 | 1 | 1 | 1 | 1 | 1 | 2 | 2 | 2 | 2 | 2 | 2 | 2 | 2 | 2 | 2 | 2 | 2 | 2 | 2 | 2 | 3 | 3 | 3 | 3 | 2 | 0 | 0 |
|  | other fruits | 263 | 203 | 166 | 165 | 164 | 164 | 163 | 164 | 166 | 167 | 168 | 169 | 170 | 171 | 172 | 173 | 174 | 173 | 173 | 173 | 173 | 169 | 175 | 180 | 193 | 216 | 209 | 238 | 244 |
|  | high vitamin C fruits | 280 | 317 | 342 | 336 | 331 | 326 | 321 | 316 | 311 | 305 | 300 | 295 | 290 | 285 | 280 | 275 | 269 | 269 | 269 | 269 | 269 | 272 | 299 | 330 | 361 | 392 | 423 | 423 | 423 |
| **Fruit juice** | 100%fruit juice | 29 | 54 | 69 | 71 | 73 | 75 | 77 | 78 | 79 | 80 | 81 | 82 | 83 | 84 | 85 | 86 | 87 | 89 | 89 | 89 | 89 | 95 | 115 | 135 | 150 | 150 | 150 | 150 | 150 |
| **Milk** | milk | 0 | 0 | 0 | 0 | 0 | 0 | 0 | 2 | 2 | 1 | 1 | 0 | 0 | 0 | 0 | 0 | 0 | 0 | 0 | 0 | 0 | 3 | 10 | 13 | 0 | 0 | 0 | 0 | 0 |
| **Nuts** | nuts | 15 | 15 | 15 | 15 | 15 | 15 | 15 | 15 | 15 | 15 | 15 | 15 | 15 | 15 | 15 | 15 | 15 | 15 | 15 | 15 | 15 | 15 | 15 | 15 | 15 | 15 | 15 | 15 | 15 |
| **Offal** | offal | 2 | 2 | 4 | 4 | 3 | 3 | 3 | 3 | 3 | 3 | 3 | 3 | 3 | 3 | 3 | 3 | 3 | 2 | 2 | 2 | 2 | 0 | 0 | 0 | 0 | 0 | 0 | 0 | 0 |
| **Other** | prepared dishes | 0 | 0 | 0 | 0 | 0 | 0 | 0 | 0 | 0 | 0 | 0 | 0 | 0 | 0 | 0 | 0 | 0 | 0 | 0 | 0 | 0 | 0 | 7 | 15 | 20 | 19 | 0 | 0 | 0 |
| **Other fat** | dressing sauces | 0 | 0 | 0 | 0 | 0 | 0 | 0 | 0 | 0 | 0 | 0 | 0 | 0 | 0 | 0 | 0 | 0 | 0 | 0 | 0 | 0 | 0 | 0 | 0 | 0 | 0 | 0 | 0 | 0 |
| **Pork** | pork (fresh) | 0 | 0 | 2 | 2 | 2 | 2 | 2 | 2 | 2 | 2 | 2 | 1 | 1 | 1 | 1 | 1 | 1 | 0 | 0 | 0 | 0 | 0 | 0 | 0 | 0 | 0 | 0 | 0 | 0 |
|  | processed meat | 0 | 0 | 0 | 0 | 0 | 0 | 0 | 0 | 0 | 0 | 0 | 0 | 0 | 0 | 0 | 0 | 0 | 0 | 0 | 0 | 0 | 0 | 0 | 0 | 0 | 0 | 0 | 0 | 0 |
|  | white jam | 0 | 0 | 0 | 0 | 0 | 0 | 0 | 0 | 0 | 0 | 0 | 0 | 0 | 0 | 0 | 0 | 0 | 0 | 0 | 0 | 0 | 0 | 0 | 0 | 0 | 3 | 9 | 14 | 9 |
| **Potatoes** | potatoes | 0 | 0 | 0 | 0 | 0 | 0 | 0 | 0 | 0 | 0 | 0 | 0 | 0 | 0 | 0 | 0 | 0 | 0 | 0 | 0 | 0 | 0 | 0 | 0 | 0 | 0 | 0 | 0 | 0 |
| **Poultry** | poultry | 11 | 40 | 51 | 50 | 48 | 47 | 45 | 44 | 43 | 42 | 41 | 40 | 39 | 38 | 37 | 35 | 34 | 36 | 36 | 36 | 36 | 39 | 45 | 51 | 61 | 75 | 91 | 117 | 121 |
| **Pulses** | pulses | 136 | 117 | 106 | 104 | 101 | 98 | 96 | 93 | 90 | 87 | 85 | 82 | 79 | 76 | 74 | 71 | 68 | 65 | 65 | 65 | 65 | 63 | 64 | 66 | 66 | 65 | 72 | 97 | 96 |
| **SFF**^2^ | croissant | 61 | 50 | 44 | 43 | 42 | 40 | 39 | 38 | 37 | 36 | 35 | 34 | 33 | 32 | 31 | 30 | 29 | 28 | 28 | 28 | 28 | 27 | 27 | 26 | 25 | 26 | 23 | 7 | 0 |
|  | sweet and fat products | 16 | 38 | 47 | 48 | 49 | 51 | 52 | 52 | 53 | 53 | 54 | 54 | 55 | 55 | 56 | 56 | 57 | 58 | 58 | 58 | 58 | 57 | 45 | 31 | 15 | 0 | 0 | 0 | 0 |
| **Snack** | Sandwiches | 0 | 0 | 0 | 0 | 0 | 0 | 0 | 0 | 0 | 0 | 0 | 0 | 0 | 0 | 0 | 0 | 0 | 0 | 0 | 0 | 0 | 1 | 9 | 17 | 23 | 28 | 34 | 34 | 49 |
|  | Salted biscuits and chips | 0 | 0 | 0 | 0 | 0 | 0 | 0 | 0 | 0 | 0 | 0 | 0 | 0 | 0 | 0 | 0 | 0 | 0 | 0 | 0 | 0 | 0 | 0 | 0 | 0 | 0 | 0 | 0 | 0 |
| **Sweet drinks** | sweet drinks | 0 | 0 | 0 | 0 | 0 | 0 | 0 | 0 | 0 | 0 | 0 | 0 | 0 | 0 | 0 | 0 | 0 | 0 | 0 | 0 | 0 | 0 | 0 | 0 | 0 | 0 | 0 | 0 | 0 |
| **Substitutes** | soya-based cheese | 2 | 4 | 5 | 5 | 5 | 5 | 5 | 5 | 5 | 5 | 5 | 5 | 5 | 5 | 5 | 5 | 4 | 5 | 5 | 5 | 5 | 5 | 5 | 5 | 5 | 5 | 5 | 5 | 5 |
|  | soya-based “dairy” products | 0 | 0 | 0 | 0 | 0 | 0 | 0 | 0 | 0 | 0 | 0 | 0 | 0 | 0 | 0 | 0 | 0 | 0 | 0 | 0 | 0 | 0 | 0 | 0 | 0 | 0 | 0 | 0 | 0 |
|  | soya-based “milk” | 73 | 16 | 0 | 0 | 0 | 0 | 0 | 0 | 0 | 0 | 0 | 0 | 0 | 0 | 0 | 0 | 0 | 0 | 0 | 0 | 0 | 0 | 0 | 0 | 0 | 0 | 0 | 0 | 0 |
|  | soya dring | 20 | 10 | 1 | 2 | 2 | 2 | 3 | 2 | 1 | 1 | 0 | 0 | 0 | 0 | 0 | 0 | 0 | 0 | 0 | 0 | 0 | 0 | 0 | 0 | 0 | 0 | 0 | 0 | 0 |
|  | plant-based substitutes | 0 | 0 | 0 | 0 | 0 | 0 | 0 | 0 | 0 | 0 | 0 | 0 | 0 | 0 | 0 | 0 | 0 | 0 | 0 | 0 | 0 | 0 | 0 | 0 | 0 | 0 | 0 | 0 | 0 |
| **Vegetable fat** | margarine | 16 | 14 | 13 | 13 | 13 | 13 | 13 | 13 | 13 | 13 | 13 | 13 | 13 | 13 | 13 | 13 | 13 | 13 | 13 | 13 | 13 | 12 | 12 | 12 | 13 | 14 | 15 | 21 | 22 |
|  | other oil | 0 | 0 | 0 | 0 | 0 | 0 | 0 | 0 | 0 | 0 | 0 | 0 | 0 | 0 | 0 | 0 | 0 | 0 | 0 | 0 | 0 | 0 | 0 | 0 | 0 | 0 | 0 | 0 | 0 |
|  | ALA-rich oil | 31 | 32 | 33 | 33 | 33 | 33 | 33 | 33 | 33 | 33 | 33 | 33 | 33 | 33 | 33 | 33 | 33 | 33 | 33 | 33 | 33 | 33 | 33 | 32 | 32 | 30 | 29 | 25 | 24 |
| **Vegetables** | soup | 0 | 0 | 0 | 0 | 0 | 0 | 0 | 0 | 0 | 0 | 0 | 0 | 0 | 0 | 0 | 0 | 0 | 0 | 0 | 0 | 0 | 0 | 0 | 0 | 0 | 0 | 0 | 0 | 0 |
|  | vegetables | 930 | 930 | 930 | 930 | 930 | 930 | 930 | 930 | 930 | 930 | 930 | 930 | 930 | 930 | 930 | 930 | 930 | 930 | 930 | 930 | 930 | 930 | 930 | 930 | 930 | 930 | 930 | 930 | 930 |
| **Wholegrain products** | whole bread | 165 | 147 | 138 | 139 | 140 | 142 | 142 | 142 | 142 | 142 | 142 | 142 | 142 | 142 | 142 | 142 | 141 | 139 | 139 | 139 | 139 | 137 | 120 | 105 | 103 | 101 | 102 | 105 | 100 |
|  | whole cereals | 50 | 31 | 19 | 20 | 20 | 20 | 20 | 20 | 21 | 21 | 22 | 22 | 22 | 23 | 23 | 23 | 24 | 23 | 23 | 23 | 23 | 21 | 17 | 13 | 14 | 16 | 13 | 0 | 22 |

Abbreviations: M, model; SFF, Sweet and fat foods

^1^M0 to M28 denote models imposing GHGe of 1.2 to 6.8 kgCO2eq/d by increments of 0.2

^2^Sweet and fat foods (SFF) include croissants, pastries, chocolate, biscuits, milky desserts, ice cream, honey and marmalade, cakes, chips, salted oilseeds, salted biscuits

### Supplemental Table 5: Consumption of food groups (g/d) in GHG-imposed scenarios - sensitivity analysis using the 95th percentile as the maximum of food group consumption)^1^

|  | M0 | M1 | M2 | M3 | M4 | M5 | M6 | M7 | M8 | M9 | M10 | M11 | M12 | M13 | M14 | M15 | M16 | M17 | M18 | M19 | M20 | M21 | M22 |
| --- | --- | --- | --- | --- | --- | --- | --- | --- | --- | --- | --- | --- | --- | --- | --- | --- | --- | --- | --- | --- | --- | --- | --- |
| Alcoholic beverages | 1 | 1 | 1 | 1 | 1 | 1 | 1 | 1 | 1 | 1 | 1 | 1 | 1 | 1 | 1 | 1 | 1 | 1 | 1 | 1 | 1 | 1 | 1 |
| Animal fat | 0 | 0 | 0 | 0 | 0 | 0 | 0 | 0 | 0 | 0 | 0 | 0 | 0 | 0 | 0 | 0 | 0 | 0 | 0 | 0 | 0 | 0 | 0 |
| Beef | 0 | 0 | 1 | 6 | 10 | 15 | 20 | 24 | 29 | 33 | 38 | 42 | 47 | 51 | 56 | 61 | 65 | 71 | 71 | 71 | 71 | 71 | 71 |
| Refined cereals | 169 | 181 | 181 | 179 | 178 | 177 | 176 | 175 | 174 | 173 | 172 | 171 | 170 | 169 | 168 | 167 | 166 | 165 | 165 | 165 | 165 | 184 | 203 |
| Dairy products | 65 | 53 | 53 | 53 | 53 | 52 | 52 | 52 | 51 | 51 | 50 | 50 | 49 | 49 | 50 | 53 | 56 | 47 | 48 | 47 | 47 | 50 | 53 |
| Eggs | 0 | 0 | 0 | 0 | 0 | 0 | 0 | 0 | 0 | 0 | 0 | 0 | 0 | 0 | 0 | 0 | 0 | 0 | 0 | 0 | 0 | 0 | 0 |
| Fish | 29 | 29 | 29 | 29 | 29 | 29 | 29 | 29 | 29 | 29 | 29 | 29 | 29 | 29 | 29 | 29 | 29 | 29 | 29 | 29 | 29 | 29 | 29 |
| Fruit | 596 | 581 | 571 | 572 | 573 | 575 | 576 | 577 | 579 | 580 | 582 | 583 | 584 | 586 | 587 | 587 | 587 | 596 | 596 | 596 | 596 | 623 | 634 |
| Fruit juice | 85 | 139 | 150 | 150 | 150 | 150 | 150 | 150 | 150 | 150 | 150 | 150 | 150 | 150 | 150 | 150 | 150 | 150 | 150 | 150 | 150 | 150 | 150 |
| Milk | 60 | 92 | 94 | 95 | 96 | 98 | 99 | 101 | 103 | 104 | 106 | 108 | 110 | 111 | 111 | 108 | 106 | 118 | 118 | 118 | 118 | 110 | 96 |
| Nuts | 15 | 15 | 15 | 15 | 15 | 15 | 15 | 15 | 15 | 15 | 15 | 15 | 15 | 15 | 15 | 15 | 15 | 15 | 15 | 15 | 15 | 15 | 15 |
| Offal | 2 | 2 | 6 | 6 | 6 | 6 | 6 | 5 | 5 | 5 | 5 | 5 | 5 | 5 | 5 | 5 | 5 | 0 | 0 | 0 | 0 | 0 | 0 |
| Mixed dishes^2^ | 0 | 0 | 0 | 0 | 0 | 0 | 0 | 0 | 0 | 0 | 0 | 0 | 0 | 0 | 0 | 0 | 0 | 0 | 0 | 0 | 0 | 0 | 0 |
| Other fat | 0 | 0 | 0 | 0 | 0 | 0 | 0 | 0 | 0 | 0 | 0 | 0 | 0 | 0 | 0 | 0 | 0 | 0 | 0 | 0 | 0 | 0 | 0 |
| Pork | 0 | 0 | 0 | 0 | 0 | 0 | 0 | 0 | 0 | 0 | 0 | 0 | 0 | 0 | 0 | 0 | 0 | 0 | 0 | 0 | 0 | 0 | 0 |
| Potatoes | 0 | 0 | 0 | 0 | 0 | 0 | 0 | 0 | 0 | 0 | 0 | 0 | 0 | 0 | 0 | 0 | 0 | 0 | 0 | 0 | 0 | 0 | 0 |
| Poultry | 4 | 39 | 44 | 42 | 41 | 40 | 38 | 37 | 35 | 34 | 32 | 31 | 29 | 28 | 27 | 25 | 24 | 27 | 27 | 27 | 27 | 30 | 38 |
| Pulses | 343 | 317 | 308 | 302 | 296 | 289 | 282 | 275 | 269 | 262 | 255 | 249 | 242 | 235 | 229 | 223 | 216 | 210 | 210 | 210 | 210 | 233 | 267 |
| SFF^3^ | 29 | 39 | 45 | 45 | 45 | 44 | 44 | 43 | 43 | 42 | 42 | 42 | 41 | 41 | 40 | 40 | 40 | 37 | 37 | 37 | 37 | 22 | 7 |
| Snack | 0 | 0 | 0 | 0 | 0 | 0 | 0 | 0 | 0 | 0 | 0 | 0 | 0 | 0 | 0 | 0 | 0 | 0 | 0 | 0 | 0 | 13 | 24 |
| Sweet drinks^4^ | 0 | 0 | 0 | 0 | 0 | 0 | 0 | 0 | 0 | 0 | 0 | 0 | 0 | 0 | 0 | 0 | 0 | 0 | 0 | 0 | 0 | 0 | 0 |
| Substitutes | 100 | 16 | 0 | 0 | 0 | 0 | 0 | 0 | 0 | 0 | 0 | 0 | 0 | 0 | 0 | 0 | 0 | 0 | 0 | 0 | 0 | 0 | 0 |
| Vegetable fat | 56 | 56 | 55 | 55 | 55 | 55 | 55 | 54 | 54 | 54 | 54 | 54 | 54 | 54 | 54 | 53 | 53 | 53 | 53 | 53 | 53 | 53 | 53 |
| Vegetables | 611 | 611 | 611 | 611 | 611 | 611 | 611 | 611 | 611 | 611 | 611 | 611 | 611 | 611 | 611 | 611 | 611 | 611 | 611 | 611 | 611 | 611 | 611 |
| Wholegrain products | 223 | 196 | 190 | 191 | 192 | 193 | 194 | 195 | 196 | 197 | 198 | 199 | 200 | 201 | 202 | 203 | 204 | 204 | 204 | 204 | 204 | 186 | 167 |

Abbreviations: M, model; SFF, Sweet and fat foods

^1^M0 to M28 denote models imposing GHGe of 1.2 to 6.8 kgCO2eq/d by increments of 0.2 under nutritional, dietary guidelines and acceptability constraints using the 95^th^ percentile as maximum of food group consumption

^2^Mixed dishes include sandwiches, dishes such as pizza, hamburger, ravioli, panini, salted pancake

^3^Sweet and fat foods (SFF) include croissants, pastries, chocolate, biscuits, milky desserts, ice cream, honey and marmalade, cakes, chips, salted oilseeds, salted biscuits

^4^Sweet drinks include fruit nectar, syrup, soda (with or without sugar)

### Supplemental Figure 1: Contribution of food groups to nutrient intake in GHG-imposed scenarios^1,2^

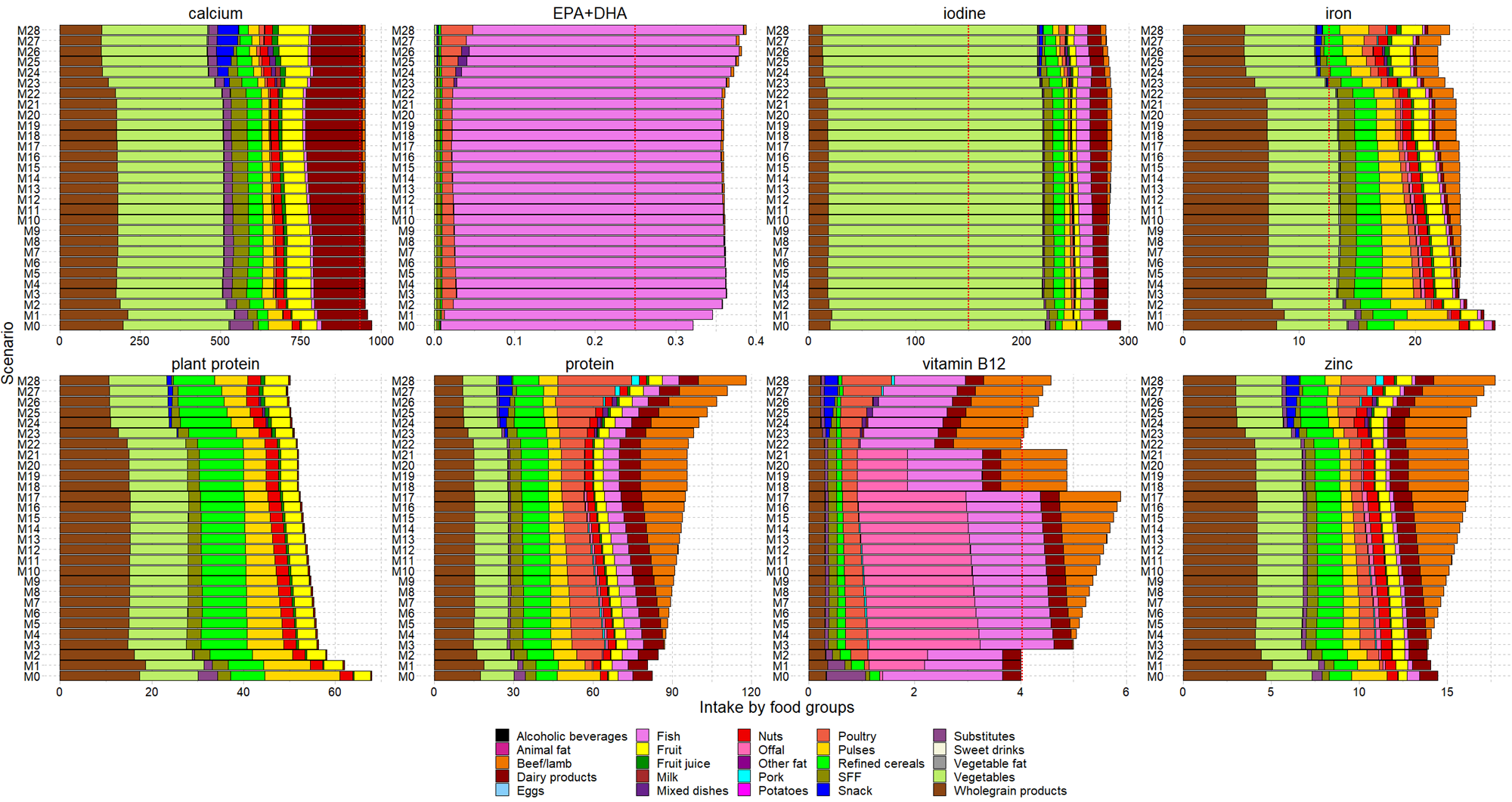

Abbreviations: DHA, docosahexaenoic acid; EI, energy intake; EPA, eicosapentaenoic acid; M:model; SFF, Sweet and fat foods

Units were mg for calcium, iron, and zinc, µg for iodine, and vitamin B12, kcal for energy intake and g for DHA, EPA and protein.

^1^Mixed dishes include sandwich, dishes such as pizza, hamburger, ravioli, panini, salted pancake, sweet and fat foods (SFF) include croissants, pastries, chocolate, biscuits, milky desserts, ice cream, honey and marmalade, cakes, chips, salted oilseeds, salted biscuits, and sweet drinks include fruit nectar, syrup, soda (with or without sugar)

^2^M0 to M28 denote models imposing GHGe of 1.2 to 6.8 kgCO2eq/d by increments of 0.2
